# A Deep Learning Approach to Forecast Short-Term COVID-19 Cases and Deaths in the US

**DOI:** 10.1101/2022.08.23.22279132

**Authors:** Hongru Du, Ensheng Dong, Hamada S. Badr, Mary E. Petrone, Nathan D. Grubaugh, Lauren M. Gardner

## Abstract

Since the US reported its first COVID-19 case on January 21, 2020, the science community has been applying various techniques to forecast incident cases and deaths. To date, providing an accurate and robust forecast at a high spatial resolution has proved challenging, even in the short term. Here we present a novel multi-stage deep learning model to forecast the number of COVID-19 cases and deaths for each US state at a weekly level for a forecast horizon of 1 to 4 weeks. The model is heavily data driven, and relies on epidemiological, mobility, survey, climate, and demographic. We further present results from a case study that incorporates SARS-CoV-2 genomic data (i.e. variant cases) to demonstrate the value of incorporating variant cases data into model forecast tools. We implement a rigorous and robust evaluation of our model – specifically we report on weekly performance over a one-year period based on multiple error metrics, and explicitly assess how our model performance varies over space, chronological time, and different outbreak phases. The proposed model is shown to consistently outperform the CDC ensemble model for all evaluation metrics in multiple spatiotemporal settings, especially for the longer-term (3 and 4 weeks ahead) forecast horizon. Our case study also highlights the potential value of virus genomic data for use in short-term forecasting to identify forthcoming surges driven by new variants. Based on our findings, the proposed forecasting framework improves upon the available forecasting tools currently used to support public health decision making with respect to COVID-19 risk.

**Research in context:** *Evidence before this study:* A systematic review of the COVID-19 forecasting and the EPIFORGE 2020 guidelines reveal the lack of consistency, reproducibility, comparability, and quality in the current COVID-19 forecasting literature. To provide an updated survey of the literature, we carried out our literature search on Google Scholar, PubMed, and *medRxi*, using the terms “Covid-19,” “SARS-CoV-2,” “coronavirus,” “short-term,” “forecasting,” and “genomic surveillance.” Although the literature includes a significant number of papers, it remains lacking with respect to rigorous model evaluation, interpretability and translation. Furthermore, while SARS-CoV-2 genomic surveillance is emerging as a vital necessity to fight COVID-19 (i.e. wastewater sampling and airport screening), to our knowledge, no published forecasting model has illustrated the value of virus genomic data for informing future outbreaks.

*Added value of this study:* We propose a multi-stage deep learning model to forecast COVID-19 cases and deaths with a horizon window of four weeks. The data driven model relies on a comprehensive set of input features, including epidemiological, mobility, behavioral survey, climate, and demographic. We present a robust evaluation framework to systematically assess the model performance over a one-year time span, and using multiple error metrics. This rigorous evaluation framework reveals how the predictive accuracy varies over chronological time, space, and outbreak phase. Further, a comparative analysis against the CDC ensemble, the best performing model in the COVID-19 ForecastHub, shows the model to consistently outperform the CDC ensemble for all evaluation metrics in multiple spatiotemporal settings, especially for the longer forecasting windows. We also conduct a feature analysis, and show that the role of explanatory features changes over time. Specifically, we note a changing role of climate variables on model performance in the latter half of the study period. Lastly, we present a case study that reveals how incorporating SARS-CoV-2 genomic surveillance data may improve forecasting accuracy compared to a model without variant cases data.

*Implications of all the available evidence:* Results from the robust evaluation analysis highlight extreme model performance variability over time and space, and suggest that forecasting models should be accompanied with specifications on the conditions under which they perform best (and worst), in order to maximize their value and utility in aiding public health decision making. The feature analysis reveals the complex and changing role of factors contributing to COVID-19 transmission over time, and suggests a possible seasonality effect of climate on COVID-19 spread, but only after August 2021. Finally, the case study highlights the added value of using genomic surveillance data in short-term epidemiological forecasting models, especially during the early stage of new variant introductions.

## Introduction

By January 31^st^, 2022, over 55 million cases and 850 thousand deaths have been attributed to SARS-CoV-2 virus in the US.^1,2^ Since the start of the pandemic, and in response to the need to allocate (often limited) resources and help guide policy making, the scientific community has sought to predict the spread of COVID-19.^3–6^ Various prospective modeling efforts exist to forecast short-term (i.e., weeks) epidemiological outcomes (cases, deaths, and hospitalizations), as well as conduct longer term (i.e., months) scenario analysis.

The approaches applied by researchers to generate short-term COVID-19 forecasts can broadly be categorized into three approaches: mechanistic, statistical, and hybrid modeling. Multiple mechanistic modeling approached have been applied to COVID-19 forecasting, which explicitly represent transmission dynamics in a population through the use of compartment models such as Susceptible-Infected-Recovered (SIR) and extensions.^7–10^ An alternative to the mechanistic approach is statistical modeling, which estimates the mathematical representation of observed behavior directly from available data. These methods typically rely upon machine learning techniques for forecasting, which most commonly include time series,^11,12^ decision tree,^13^ and deep learning approaches.^14,15^ The long short-term memory network (LSTM) occupies an important position among all deep learning methods due to its advantages in processing time series data. Researchers have applied various frameworks of LSTM to forecast COVID-19 epidemiological outcomes for the U.S. at different spatial resolutions.^16–19^ The third modeling approach merges mechanistic and statistical methodologies, here referred to as hybrid models, which take advantage of the strengths of each method to improve model performance.^20^ For example, the DeepGLEAM model combines a stochastic compartmental simulation model with deep learning for COVID-19 forecasting.^21^ All approaches utilized to date have their own strengths and weaknesses. Mechanistic models are good at providing epidemiological explanations for observed behavior, and are capable of explicitly analyzing different policies such as mask mandate and other social distancing measures through model parameterization; however, these modeling frameworks are limited in their ability to capture rapid changes in disease spreading behavior or consider potential risk factors other than those represented within the compartmental framework.^16^ In contrast, statistical models, while flexible enough to include any potential variable of interest, heavily rely on the quality and availability of the required input data, and critically, the outputs are not constrained to adhere to feasible viral dynamics. One approach to mitigate the method-specific weaknesses is to use ensemble models, such as the CDC COVID-19 Forecast Hub model, which compile multiple models of various approaches within a single prediction framework.^22^ This approach has consistently proven to be the most robust, and best performing approach for short term COVID-19 forecasting efforts, and thus why we evaluate our model against it.

Whatever the method, a recognized shortcoming in the existing COVID-19 modeling literature is the lack of rigorous and robust evaluation, which is critical to assess and compare model performance.^23^ On October 19^th^ 2021, the CDC COVID-19 Forecast Hub published the EPIFORGE guidelines to attempt to improve the quality of models, highlighting the importance of consistency, interpretability, reproducibility, and comparability of models.^24^ However, most model evaluation presented in the published literature remains incomprehensive.^23^ Many models are evaluated for a single forecasting period, according to a single error metric, and sometimes not evaluated retrospectively at all.^23^ Furthermore, many of the existing studies do not account for critical factors, such as human behavior, which are available through mobility data and/or real-time survey data. Additionally, there is a substantial gap between model development and model implementation for real-time forecasting, and many of the models mentioned above lack guidance on when and where each model would be most suitable, let alone information on if, when and where they were applied.

In this study we address these existing gaps in the literature and provide a more reliable source of COVID-19 forecasts for policymakers and the public. We proposed a deep learning model to forecast the US COVID-19 cases and deaths at the state level for 1- to 4-week forecasting windows. The model incorporates epidemiological (cases, deaths, hospitalizations, vaccinations), mobility, survey, climate, demographic, and virus genomic data. We assess the model performance based on multiple error metrics, as well as for varying time periods, regions, and as a function of different outbreak phases, namely periods of intense growth, decline and stability. Lastly, we implement a retrospective case study incorporating SARS-CoV-2 genomic surveillance data to demonstrate the value of implementing new variant introductions into forecasting tools.

## Data

The proposed LSTM model is heavily data driven and trained using multiple disparate categories: epidemiological, mobility, survey, climate, demographic, and virus genomic data. The time-varying data are available at a daily resolution for each US state. We rely on a combination of raw and derived metrics as inputs, which are listed in Table 1, and each is described in detail in Appendix Section 1.

**Table 1:** Summary of input data

| State-Level Data | Data Processing | Data Smoothing | Sources |
| --- | --- | --- | --- |
| ***Epidemiological data*** |  |  |  |
| COVID-19 cases/deaths | Raw | 7-day moving average | 1 |
| Growth rate of cases/deaths | Derived | 7-day moving average | 1 |
| Vaccination coverage | Raw | 7-day moving average | 15,17 |
| Hospitalization data | Raw | 7-day moving average | 30,31 |
| ***Mobility data*** |  |  |  |
| Importation risk | Derived | 7-day moving average | 1,32 |
| Mobility ratio | Derived | 7-day moving average | 32 |
| Visits ratio for 21 different destinations | Derived | Principal component analysis | 32 |
| ***Survey data*** |  |  |  |
| COVID-like symptoms in community | Raw | Raw data has already been smoothed | 31 |
| ***Climate data*** |  |  |  |
| Temperature (°C) | Raw | 7-day moving average | 33 |
| Precipitation (mm/day) | Raw | 7-day moving average | 33 |
| ***Demographic data*** |  |  |  |
| Population | Raw | - | 34 |
| Proportion of people over 65 | Raw | - | 34 |
| ***Virus Genomic data*** |  |  |  |
| Variant cases | Derived | 7-day moving average | 35 |

## Methodology

COVID-19 transmission patterns have proven complex over time. Thus, forecasting even near-term disease dynamics requires a robust predictive modeling framework and carefully selected input data streams. Critically, the framework must account for nonlinear interactions between the considered factors affecting the transmission dynamics and uncertainty in their time-dependent impact on observed transmission dynamics. We therefore propose a multi-stage deep learning framework, which, at each stage, forecasts a chosen target variable for the seven days ahead (e.g., one-week ahead forecast). The multi-stage model builds off the initial first stage prediction to forecast an additional week out and continues to implement this iterative approach one stage at a time, to predict further into the future. In this paper, we will focus on 4-stage forecasting, which generates 4-week ahead predictions, consistent with the CDC COVID-19 Forecast Hub.^25,26^ However, the framework can be applied to shorter- and longer-term horizons.

### Multi-Stage LSTM Network Architecture

The multi-stage framework consists of two neural network branches, connected in parallel, as illustrated in Figure 1. The main branch (main model) predicts the target epidemiological variables of interest, while the secondary branch (feature model) predicts independent features to populate the data streams used as input in the main model. The target variable for the main model is either weekly incident cases or weekly mortality rate; for the features model, target variables are all other independent time-varying features that serve as predictors for the main model. An example of a model output is shown in Figure 1.C, for New York state, specifically, the forecasted weekly cases for each of the four weeks following October 17^th^, 2020. Additional implementation of the multi-stage framework, details of model formulations, and model parameterization are described in detail in Appendix Section 2·1 to 2·3.

**Figure. 1.**
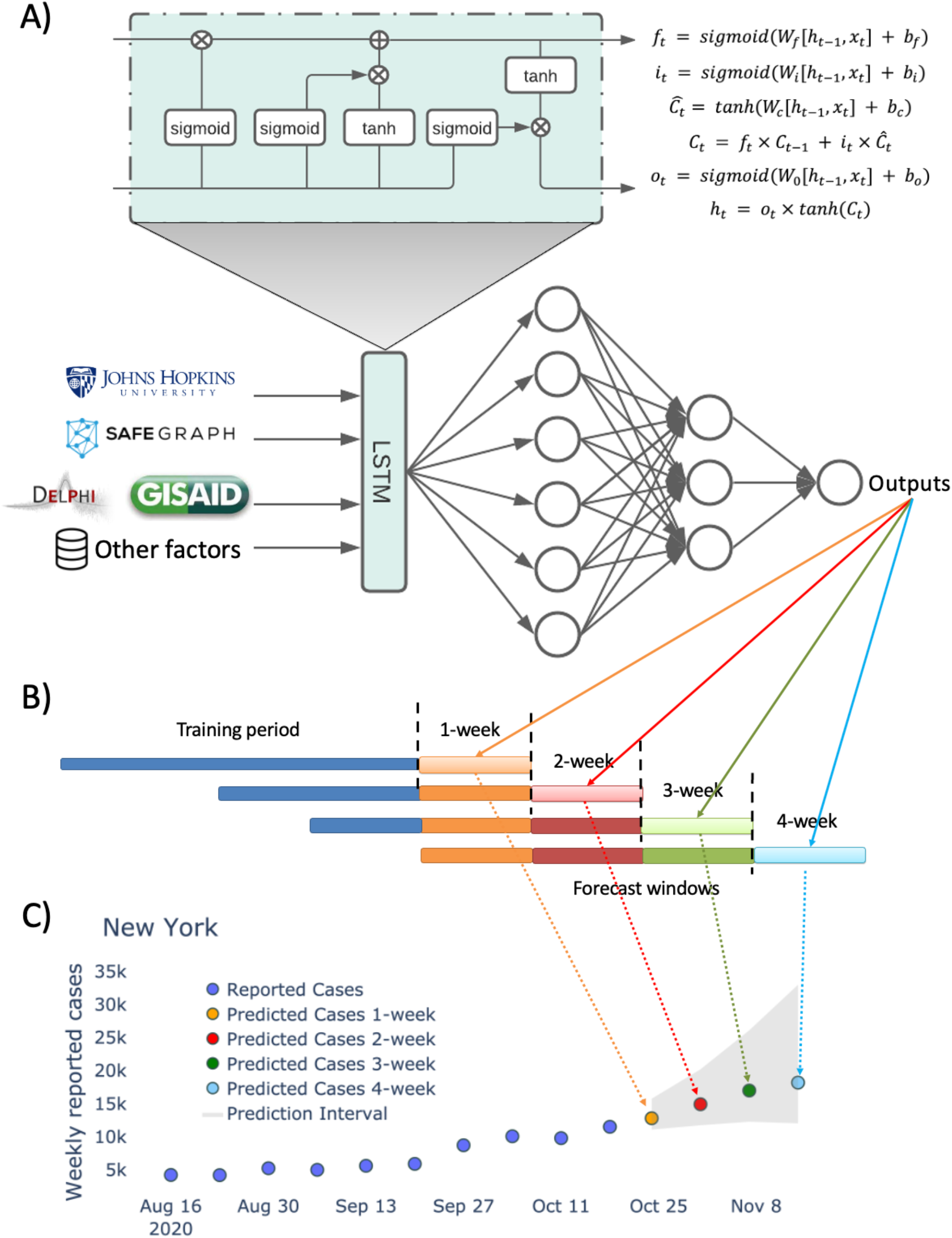
A) Network architecture of the multi-stage LSTM model. B) Prediction structure of the multi-stage LSTM model. At the initial stage, the model uses the most recent data as input, then at the later stage, the model adapts previous prediction as input to make further predictions. The transparent colors represent the model’s output, and solid colors represents the model’s inputs. C) An example forecasting of the multi-stage LSTM model.

### Model Evaluation

We conduct a robust evaluation of the model performance, explicitly assessing its performance as a function of space, time, and outbreak phase. All assessment is conducted over a long horizon (52 weeks, spanning all epidemiological weeks from August 2020 to August 2021), and evaluated using three different error metrics: a) Absolute Error (AE), b) Percentage Absolute Error (PAE), and c) Weighted Interval Scores (WIS).^20^ The definition of each error metric is described in Appendix Section 2·4. The first two metrics measure the accuracy of point predictions, while the last metric is intended to evaluate the model predictions as a probability distribution. For all experiments, we use JHU CSSE actual weekly reported cases and deaths ^1^ as the ground truth data to compute the error metrics. While this analysis is retrospective, the evaluation is based on data that would have been available at the time of prediction, to align with the real-time forecasting constraints. For space constraints, the PAE results are presented throughout this section, and the WIS and AE results, when relevant, are provided in relevant sections throughout the Appendix. We compare our results to the CDC ensemble model,^20^ which we use as the benchmark because it has consistently proven to be the top performing model in the CDC COVID-19 Forecast Hub,^22^ among dozens of individually contributed models (ensemble members).

We also conduct sensitivity analysis to assess the contribution of each variable to the model performance, by evaluating different combinations of input features (Appendix Section 2·5). Due to time constraints and computational cost, the sensitivity analysis only applies to PAE and AE.

## Results

Results for the LSTM model forecasted cases for 1-, 2-, 3- and 4-week forecasting windows, for every state in the US are presented in this section. Equivalent results for deaths forecasts are described in Appendix Section 3·8. We present our model performance as a function of time, space and different outbreak phases. We then conclude this section with results from a case study that supplements the input data streams with variant cases from available SARS-CoV-2 genomic surveillance data. The case study is conducted for a subset of states with the highest quality virus genomic data, and the 2021 summer period, to align with the delta wave in the US. In Appendix Section 2·5 we present results from a sensitivity analysis conducted to assess the contribution of each variable in prediction.

### Model Performance Across Time

Figure 2 illustrates the relative performance of the LSTM against the CDC ensemble model for each of the 52-week periods evaluated, for 1 to 4 week forecast windows, highlighting the performance variability over time. Each pair of bar plots represents PAE distribution for all the states at a given week, where the green bar represents the error distribution for the multi-stage LSTM model, and the yellow bar represents the error distribution for the CDC ensemble model. The red curve represents the weekly reported cases at the national level. The left y-axis represents the PAE by different forecasting windows and right y-axis represents national level reported cases.

**Figure. 2.**
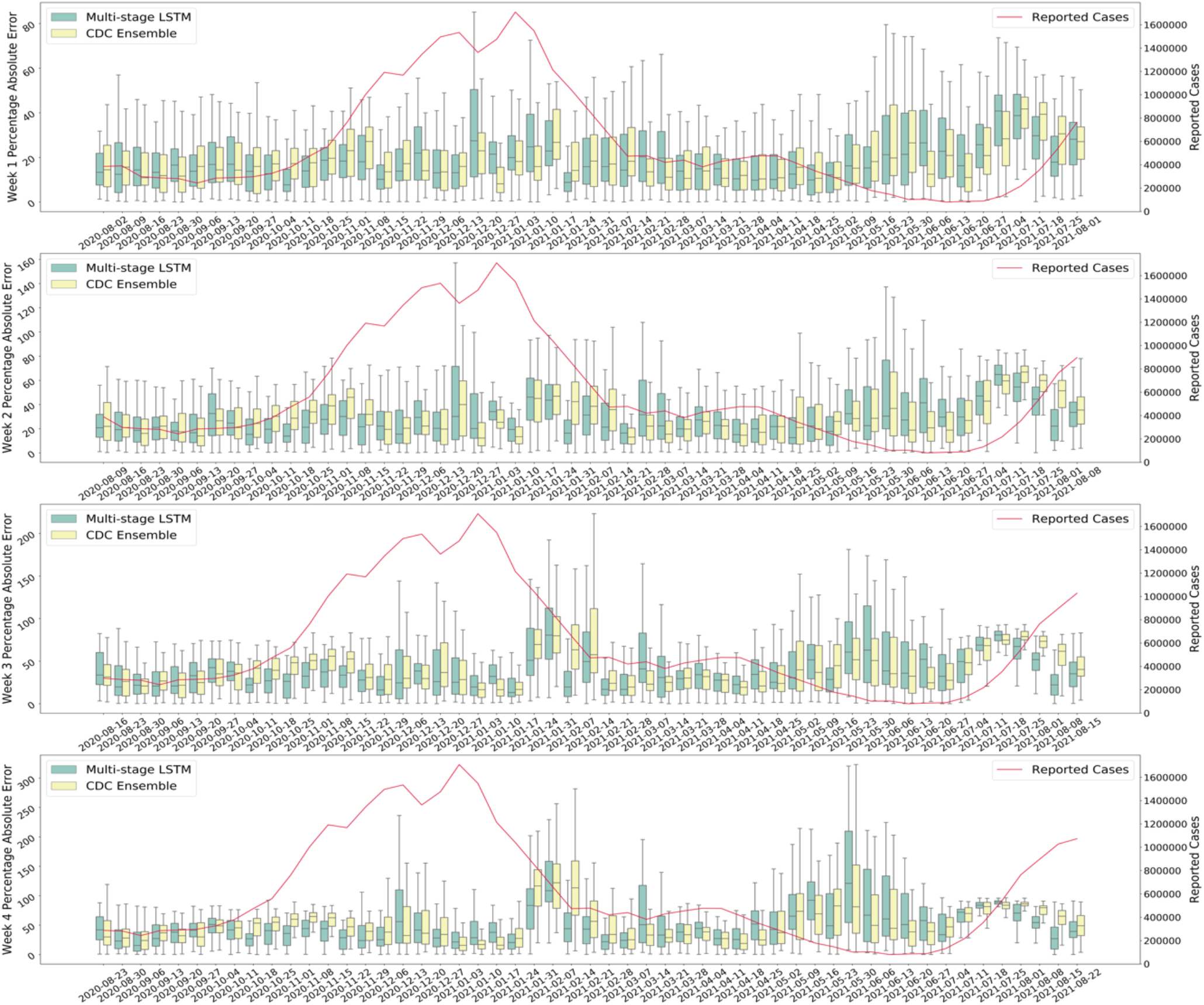
Comparison of model performance between the multi-stage LSTM Model and the CDC ensemble model based on PAE. Each pair of bar plots represents PAE distribution for all the states at a given week, where the green bar represents the error distribution for the multi-stage LSTM model, and the yellow bar represents the error distribution for the CDC ensemble model. The red curve represents the weekly reported cases at the national level. The left y-axis represents the PAE by different forecasting windows and right y-axis represents national level reported cases.

For the time period evaluated the model consistently outperforms the CDC ensemble, especially during case surges, and for longer (3 and 4 weeks ahead) forecast windows. The average PAE across all states and weeks is 22%, 32%, 44% and 57% for the 1 to 4 week forecast windows, respectively. As the forecasting window increases, the variability in performance across states further increases, as indicated by the wider bars. Figure 2 also reveals how the model performance varies with respect to the different waves of the pandemic. The model performance is relatively stable for the first five months of the study period (August 2020 to November 2020), but much more variable in performance in January 2021 and May 2021, which both correspond to periods when the cases transitioned from decreasing to more stable rates. The results for WIS and AE reveal consistent performance patterns, as illustrated in Appendix Section 3·1.

### Model Performance Across States

Figure 3 illustrates the average performance over all 52 weeks, for each state, highlighting the performance variability across space. The color scales represent the magnitude of the error metric; the scales of PAE are fixed in 10–90 range. The deeper color corresponds to larger error. Equivalent evaluations for AE and WIS are included in Appendix Section 3·2. While there are no clear spatial patterns of model performance for 1-week ahead forecast, a spatial pattern becomes evident as the forecast window increases. For the 2 to 4-week forecast windows, the PAE is relatively larger for midwestern states and smaller for southeastern states. Reasons for this are addressed in the discussion section.

**Figure. 3.**
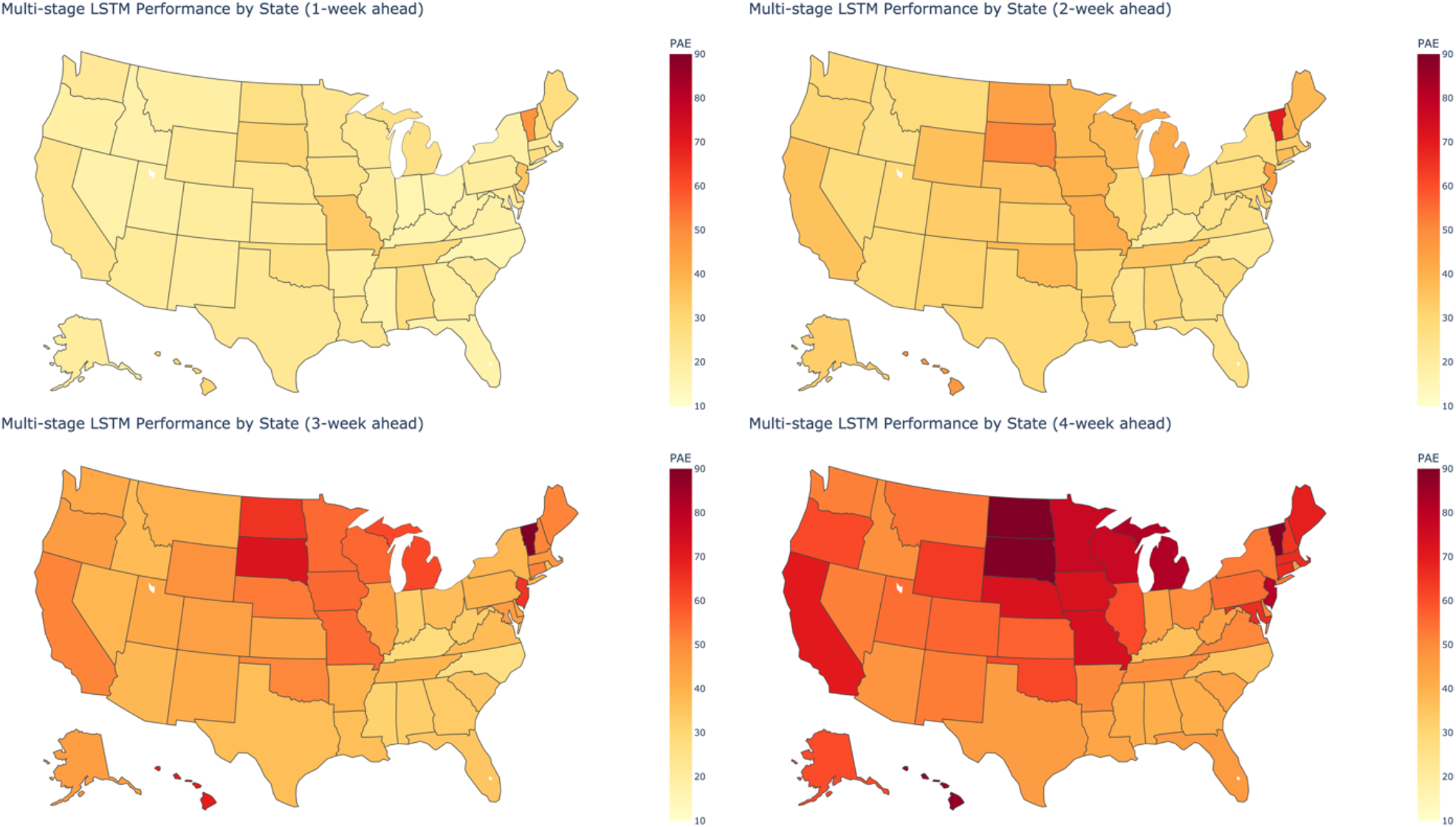
State-specific average model performance based on PAE (over all epidemiological weeks) for varying prediction windows of one- to four-week out predictions. The color scales represent the magnitude of the error metric; the scales of PAE are fixed in 10–90 range. The deeper color corresponds to larger error.

### Model Performance by Outbreak Phase

In addition to examining performance variability over fixed space and time, we also evaluate the model performance as a function of the outbreak phase. To do this, we generate five outbreak phases based on the weekly average incidence growth rates and assign each state-week pair accordingly. We apply 5-quantiles clustering according to the relative magnitude of growth rate, the five groups are classified as: 1) fast increasing (growth rate above 0·017); 2) slightly increasing (growth rate between 0·005 and 0·017); 3) flat (growth rate between -0·004 and 0·004); 4) slightly decreasing (growth rate between −0·016 and −0·004); and 5) fast decreasing (growth rate below - 0·016). The assignment of the weeks to categories is presented in Appendix Figure 15. After the phase category assignment, we evaluate the performance for all state-week pairs in each of the five phase groups independently.

Figure 4 shows the model performance of the multi-stage LSTM model by different outbreak phases, the colors represent different outbreak phases, and each bar represents the distribution of PAE in corresponding outbreak phases. This result reveals that the model performs best in the stable period and has the highest variability when cases change rapidly, consistent with the same evaluation for the CDC Ensemble model (Appendix Figure 16). Equivalent evaluation based on WIS are shown in Appendix Figure 17 and 18. In addition to evaluating the LSTM and CDC Ensemble model separately, we also compare both models under each outbreak phase (see Appendix Section 3·6). As shown in Appendix Figure 19 and 20, when growth is classified as *fast increasing*, the multi-stage LSTM model outperform the CDC ensemble model over 60% of the time for all forecast windows. For the *slightly increasing* and *fast decreasing* periods, our model slightly outperforms the CDC ensemble. However, the performance of the model is lower than the CDC ensemble during periods of outbreak stability and slight declines (e.g., December 2020 and May 2021).

**Figure. 4.**
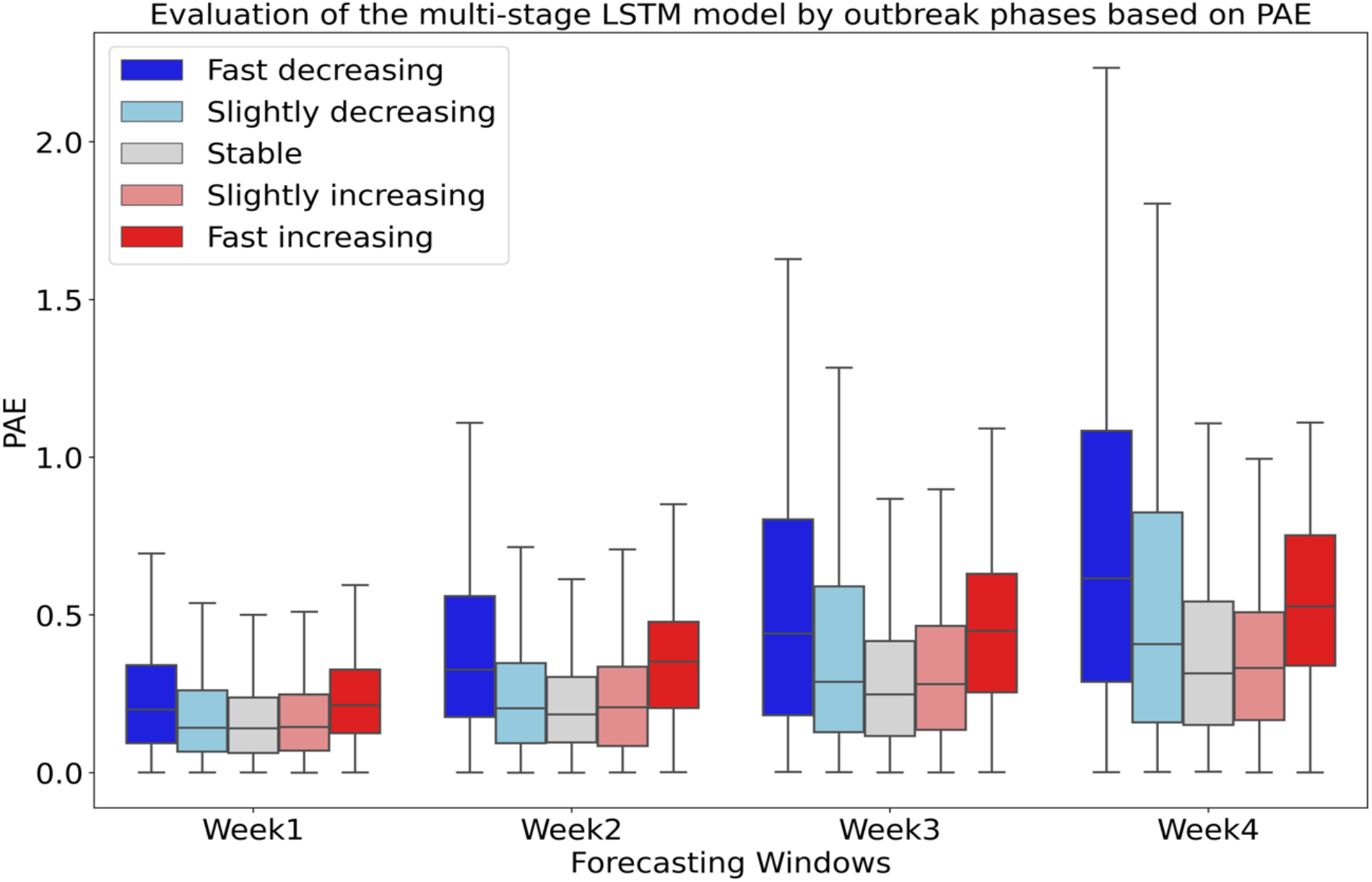
Evaluation of the multi-stage LSTM model by outbreak phases based on PAE. The colors represent different outbreak phases, and each bar represents the distribution of PAE in corresponding outbreak phases.

### Case Study with SARS-CoV-2 Genomic Surveillance Data

The US has experienced multiple waves of incident cases, often driven by new variants. In this case study, we conduct a retrospective analysis to explore the value of including variant cases from available SARS-CoV-2 genomic surveillance data in improving COVID-19 outbreak prediction using our proposed modeling framework, based on the hypothesis that genomic data may act as a signal for forthcoming changes in transmission patterns and therefore help improve prediction accuracy.^27^ Here we focus on forecasting state-level confirmed cases in the US between June 1 and August 31, 2021, capturing the wave caused by the Delta variant. We implement the analysis for the 39 selected states that sequenced at least 5% of reported cases from May 1 to August 31, 2021. We generate new variant-specific case time series (as the product of the daily proportion and total daily cases reported), which are used as inputs in the model. Details of the virus genomic data preprocessing are documented in Appendix Section 1·6. We select the top three variants with the highest proportion during June and September 2021 as new variant-specific time series, i.e., Delta, Gamma, and Alpha. In addition, we also create a fourth time series (“other”) representing the sum of all other circulating SARS-CoV-2 lineages. The inclusion of “other” category enables us to capture the introduction of new variants, in addition to other known circulating variants. When applying the model, the selection of the variant-specific time series can be adjusted dynamically, based on the most recent data.

Figure 5 illustrates the results for three different models: (a) Multi-stage LSTM model without variant cases data, (b) Multi-stage LSTM model with variant cases data and (c) CDC Ensemble model. The x-axis is the week that the predictions are made on. Each pair of bar plots represents PAE distribution for the selected states at a given week, where the green bar represents the error distribution for the multi-stage LSTM model without variant cases data, purple bar represents the error distribution for the multi-stage LSTM model with genomic data, and the yellow bar represents the error distribution for the CDC ensemble model. Results from the case study suggests that the inclusion of variant cases data have varying levels of impact, dependent on the time period. Specific trends are noted in the discussion section. The results based on AE and WIS are shown in Appendix Section 3·7.

**Figure. 5.**
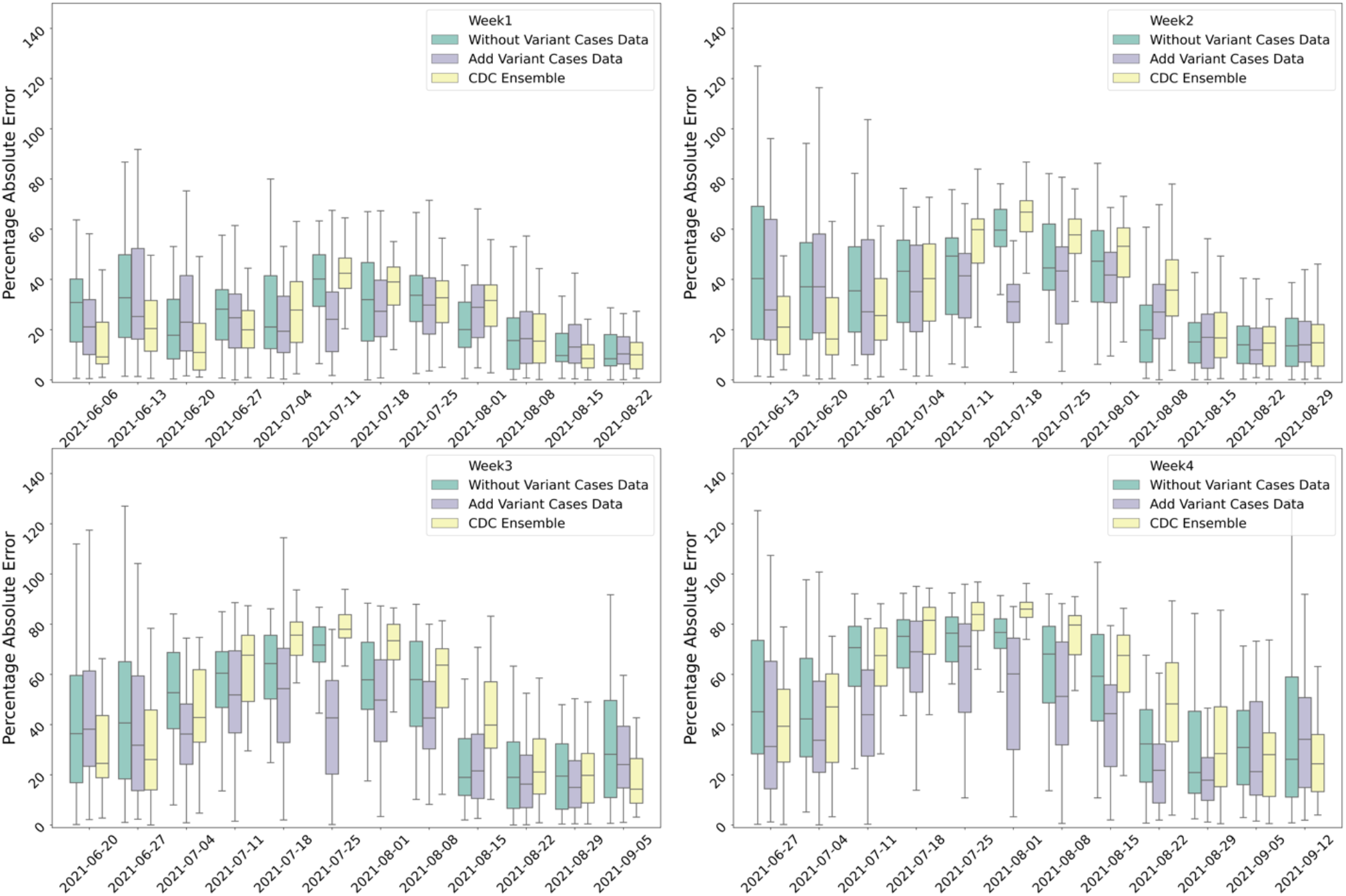
Model performance based on PAE for three different models: (a) Multi-stage LSTM model without variant cases data, (b) Multi-stage LSTM model with variant cases data and (c) CDC Ensemble model. The x-axis is the week that the predictions are made on. Each pair of bar plots represents PAE distribution for the selected states at a given week, where the green bar represents the error distribution for the multi-stage LSTM model without genomic data, purple bar represents the error distribution for the multi-stage LSTM model with genomic data, and the yellow bar represents the error distribution for the CDC ensemble model.

Notably, this study is retrospective, and therefore is not subject to the real-time reporting limitations of SARS-CoV-2 genomic data from sequences COVID-19 cases. Specifically, the average time lag in genomic data reporting is 26 days,^28^ whereas we assume data is available with a seven day lag. While not feasible at present, this study highlights the potential value of timely and open virus genomic surveillance as a pandemic forecasting tool.

### Model Selection

We conduct sensitivity analysis to assess the importance and contribution of various input features and training periods to identify the best performing model. We assign features into four categories (epidemiological, mobility, survey, and climate data). The complete set of features considered, and category assignment are listed in Table 1. Four models are constructed which include different combinations of available features, namely 1) a simple basis model with only epidemiological data, 2) a model with epidemiological and mobility data, 3) a model with epidemiological, mobility and survey data, and 4) a model with all features. We further conduct the equivalent model comparison for two discrete time periods aligning with pre and post available vaccines, specifically divided on February 1, 2021, approximately when vaccination roll out began in the US. The results comparing the performance of these four models for the entire period and two discrete periods are shown in Appendix Figures 7, and 8, respectively. The results reveal that the model with epidemiological, mobility, and survey data has the best overall performance. However, the contribution of each input feature can vary across time; this is expanded upon in the discussion section. Finally, the analysis performed for COVID-19 deaths as a response variable is presented in Appendix Figure 26, where model 3) and 4) have similar performance. Additional sensitivity analysis on model’s input parameters is included in the Appendix Section 2·5.

## Discussion

### Spatiotemporal Variability of Model Performance

Our analysis reveals a high variability in model performance as a function of the forecast window, chronological time and space. The performance over the 52 weeks evaluated is closely tied to the observed outbreak dynamics, and figure 2 highlights the impact of rapidly changing dynamics on the model performance. The model performs worse around the inflection period (especially when cases’ trend changes from decreasing to stable), and gradually improves as case (and death) rates stabilize. In terms of spatial patterns, for the time period evaluated model is more accurate in eastern and southeastern states, compared with midwestern states. This pattern is further confirmed by comparing the model performance with the CDC ensemble model. This spatial pattern can be partially explained by the difference in case trends across these regions. Specifically, during October 2020 to December 2020, midwestern states experienced the fall COVID-19 wave ahead of most of the country. Specifically, midwestern states started to show a decreasing trend while cases were increasing elsewhere (see Appendix Section 3·3). Because the model is trained using the data for all states for each prediction period, the predictions will be guided by the most dominant trend, and the model may underperform for any states not experiencing the same patterns. As an extension of this work, one could develop group-specific models through a cluster-based training setup or a more deliberate design of loss function, and as such, generate forecasts for each sub-group. Additionally, as expected, the model performance decreases as the forecasting window increases. This outcome is partially an artifact of the multi-stage nature of the modeling framework, which is sensitive to accumulative uncertainty in the input data and error propagation in the model outputs; *e*.*g*., predictions generated for each week are used as inputs for the following week’s prediction. Therefore, in periods of high instability, the one-week ahead predictions can be more erroneous, thus the error will be larger for longer forecast windows relative to the same forecast window in more stable periods. Overall, the observed spatial and temporal variability in model performance highlights the importance of identifying and communicating the optimal performance conditions for a given model before it is shared publicly or relied upon by decision makers.

### Model Performance Varies by Outbreak Phase

In Figure 2, the LSTM model is shown to perform consistently better than the CDC ensemble model in the periods of rapid outbreak growth (e.g., October 2020 to November 2020, July 2021) and decline (e.g., January 2021). To further explore model applicability, we evaluated model performance as a function of the outbreak phase, namely periods of growth, decline or stability, which were designated by five discrete categories. For the nine most populated states, most of the weeks in fall 2020 and summer 2021 are assigned to either fast or slightly increasing phase categories (Appendix Figure 18). The results highlighted in Figure 4 reveal the LSTM model to perform best in stable periods, and poorest in periods of extreme growth and decline. However, critically, the comparison of our LSTM model against the CDC Ensemble as a function of the outbreak phase, presented Appendix Figure 19 and 20, reveals that the multi-stage LSTM model performs relative better during the most critical phases of fast growth and fast decreases. This variation in forecasting accuracy during the rapidly changing outbreak phases is consistent with COVID-19 forecasting literature.^22^ Future work should consider relaxing continuous forecasting outputs, and focusing on categorical predictions, which may be able to be generated more accurately and reliably. Our analysis also highlights that model selection should consider model performance relative to the phase of the outbreak, in addition to the fixed time and location the model is applied to.

### Model Evaluation Is Sensitive to Performance Metric Chosen

A major focus of this analysis is to explore the how model performance relates to the metrics chosen for evaluation. As illustrated in the Appendix Section 3·1, the performance of the LSTM and CDC ensemble model can vary significantly, dependent on the error metric selected. This occurs due to the way the metrics are mathematically defined (Appendix Section 2·4), in particular, whether they are normalized to account for potentially large variations in the magnitude of the predictor variable or not, as well as how they account for uncertainty bounds. For example, AE has a positive correlation with confirmed case counts, therefore the states and outbreak periods with the highest reported case values will have higher AE scores; this is the case for California, New York, and Florida (Appendix Figure 12). In contrast, PAE is normalized by case levels, and is therefore more likely to have a higher relative value when case rates are low because small variabilities in the estimated versus observed incidence rate will be amplified. This behavior is illustrated during summer 2021 in states with lower populations like Maine, New Hampshire, and Vermont, when the weekly confirmed cases are below 50 (Figure 3). For all forecasting windows, the results are shown to be sensitive to the error metric chosen, and critically, the selection of the best performing model for a given state is dependent on the metric chosen for evaluation. However, as the forecasting window increases, the LSTM model appears to consistently outperform the CDC ensemble model for the southeastern states (i.e., Virginia, North Carolina, South Carolina) according to all metrics. This analysis highlights the need to consider multiple metrics in evaluating models, in order to improve model selection and robustly assess model performance.

### Model Sensitivity to Input Data Streams

Results from the sensitivity analysis to assess the importance and contribution of various input features revealed the best performing model included all the features except climate data. Our analysis reveals that a model solely reliant on epidemiological data performed worst, while adding mobility and survey data reliably improved model performance, especially for longer forecasting windows. These results support the inclusion of preprocessed mobility variables and real-time survey variables in learning model frameworks such as the proposed LSTM model. While the epidemiologic, survey and mobility variables revealed similar roles across the entire study period, and each of the separate periods evaluated, the role of climate variables is less clear. The inclusion of climate variables did not initially appear to improve predictive capability (when considered across the entire study period), however, when we divided the study period into two discrete periods, the role of the climate data changed. For the period between August 2020 and February 2021, the inclusion of climate data did not improve the model performance, however during the second phase of the study period, between February and August 2021, the inclusion of climate variables increased the model performance (Appendix Figure 8). These results suggest a differing role of climate on COVID-19 transmission in the first and second year of the pandemic, which aligns with other literature.29 We hypothesize in the first year of the pandemic factors other than climate, such as behavior and underlying population immunity, dominated the role of climate, and/or the role of climate is being captured indirectly through other predictors (e.g., higher temperatures lead to behavioral changes which can be captured through the survey and mobility data sets). While this preliminary analysis sheds some light on the possible role of climate and seasonality of COVID-19, this is an area in need of further research.

### Inclusion of SARS-CoV-2 Genomic Surveillance Data Improves Model Performance

The case study, designed to capture new variant introductions and variant growth rates, highlights the value of using SARS-CoV-2 genomic surveillance in short-term epidemiological forecasting, specifically with regards to early identification of inflection points. The inclusion of genomic data consistently improved the model performance within two weeks after the average proportion of Delta variant above 15% for most of the 39 states included in the cases study. Specifically, for predictions between epidemiological weeks June 20, and July 25, 2021, the LSTM model with variant cases data performed better than both the reference LSTM model (without the variant cases data) and the CDC ensemble model, especially for the longer three- and four-week forecasting windows. This is approximately the period when there is a switching of dominated variant between Delta variant and Alpha variant (Appendix Figure 6). However, this performance ranking changed immediately after the Delta variant proportion reached 100%. A possible explanation for this is that when the Delta variant proportion reached 100%, the proportion of Gamma, Alpha, and other variant specific cases suddenly dropped to zero, and our LSTM model requires a learning period to adapt to this change in the input data stream. As evidence for this hypothesis, the performance of the two LSTM models (with and without variant cases data) converges a few weeks after the Delta variant reaches 100% (Figure 5).

### Limitations

There are several limitations to this study, primarily resulting from data issues, and imposed methodological constraints. Most critically, there are challenges posed by the quality and availability of the data relied upon, for both the health outcomes data sets used to represent ground truth, as well as the input data streams. Given the intended real time use of this framework, the best available data at the time of generating the forecast were used to both train and evaluate the model, and as such, unresolved anomalies, biases and inaccuracies in the data directly affect performance. Further data quality issues such as spatiotemporal biases, sample size and data gaps also posed challenges, and were more prevalent in the data sets used to capture human behavior, e.g., survey data. In addition to quality of the data used, certain critical features are excluded from the model, such as government policies and policy compliance rates, as well as other behavioral data. Future work should explore the inclusion of these additional data sources to further enhance model performance. In addition to data issues, the LSTM model is fully empirical, i.e., it does not have a mechanistic component, therefore the actual infection dynamics are not constrained by feasible outbreak scenarios, which can result in unrealistic predictions. The empirical nature of the model also constrains the forecasts to previously observed transmission patterns (within the training time window); thus, the model will perform poorly when the transmission dynamics dramatically differ (exceed) from prior behavior.

## Conclusion

We introduced a flexible deep learning framework that utilizes a broad set of data types (epidemiological, mobility, survey, climate, demographic, and virus genomic) to forecast COVID-19 cases and deaths in real time. The novel multi-stage forecasting routine uses an iterative approach, building on one stage’s outputs to generate the next stage’s predictions. We applied our framework for the United States at a weekly temporal resolution and state-level spatial resolution, for a four-week planning horizon. We evaluated our model at each epidemiological week over the 52-week period between August 2020 to August 2021, and quantified performance using three different error metrics. We further break down the performance as a function of outbreak phases, location, time, and forecasting window. While the model is shown to perform well in multiple settings, the results from this analysis illustrate a variable performance of the model across the considered dimensions. This variability is driven by the complex, uncertain and evolving role of the critical contributing factors that drive COVID-19 transmission dynamics. This includes, for example, changes behavior, immunity, climate, the environment, and viral dynamics. Based on these findings, forecasting models should be accompanied with specifications on the conditions under which models performs best (and worst), in order to maximize their value and utility in aiding public health decision making. Extensions of this work include applying it at higher spatial resolutions (e.g., at the county level), and for predicting other response variables (e.g., hospitalization rates). Further, we selected a simple LSTM as the model’s building block since it is a state-of-art framework for processing time dependent data, however, rigorous inter-comparisons with other deep learning techniques should be conducted.

## Data Availability

All data produced in the present study are available upon reasonable request to the authors.

## Author Contributions

LG, HD, and ED contributed to the conceptualization and design of the study. HD and ED collected the data and conducted the analysis. HD led the writing of the original draft. All authors edited the manuscript, discussed results, and provided feedback regarding the manuscript. LG supervised the study and acquired funding. HD and ED have verified the underlying data. All authors had full access to the data and approved the manuscript for publication.

## Acknowledgements

This work was funded the NSF Rapid Response Research (RAPID) grant Award ID 2108526 and the CDC Contract #75D30120C09570.

## Supplementary Appendix

### 1. Data and Preprocessing

The proposed LSTM model is trained using multiple disparate categories of data including epidemiological, mobility, survey, climate, vaccine coverage, demographic, and genomic data. The time-varying data are all available at a daily resolution, and state spatial resolution. We use a mixture of preexisting and generated metrics as input; all the variables and their corresponding categories are summarized in the table 1 in the main manuscript, and described in detail below:

#### 1.1 Epidemiological data

Previous COVID-19 modeling studies have relied upon a wide range of data types, with epidemiological data being the most central to the efforts. Potential epidemiological variables include reported cases and deaths, unreported or undetected infections and fatality, incidence rate, mortality rate, case-fatality ratio, growth rates, testing data, vaccination coverage, and hospitalization data. ^1–3^

##### 1.1.1 Cases and Deaths

Our study utilizes the county-level, daily reported COVID-19 case, death and vaccination data ranging from April 1, 2020 to August 31, 2021 as its primary epidemiological inputs. The data is sourced from the Center for Systems Science and Engineering (CSSE) at Johns Hopkins University. ^4^ This dataset serves as the gold standard for reliable and official reported state- and county-levels cases and deaths for the US. The start date of May 1, 2020 was chosen to minimize the possible effect of underreporting at the early stages of the pandemic. The raw case and death data are aggregated to the state level. A 7-day moving average is used to address noise due to reporting issues and variable day-of-week patterns.

##### 1.1.2 Case and Death Growth Rate

The smoothed timeseries are also used to derive additional epidemiologic parameters used as latent variables in our modeling framework, namely growth rates and incidence rates. The growth rates (GR) for cases and deaths are calculated as follows:

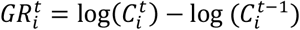

Where 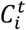 represents the smoothed cases or deaths for state *i* on day *t*. The case and death incidence rates are computed by normalizing the data by population, to generate daily cases and deaths per 100,000 persons, for each state.

##### 1.1.3 Vaccination data

Vaccine induced immunity is considered to be an essential strategy for reducing COVID-19 harm. In our model we utilize state-level vaccination data from Johns Hopkins CRC,^5^ which is collected from the US CDC Vaccine Tracker ^6^ and local health agencies. We adopt its daily state-level complete vaccination data normalized by population as one of the inputs.

##### 1.1.4 Hospitalizations data

The U.S. Department of Health & Human Service (HHS) publishes datasets “COVID-19 Reported Patient Impact and Hospital Capacity by State” via healthdata.gov. ^7^ The original dataset contains multiple columns that break the patient and hospital resources into several categories. We use cleaned COVID-19 hospitalization provided by the Delphi group at Carnegie Mellon University API. ^8^ We used 7-day moving average smoothed “inpatient_bed_used_covid” time series for our death’s prediction model.

#### 1.2 Mobility derived metrics

Previous studies have shown that aggregate human mobility patterns can be used to evaluate the impact of certain non-pharmaceutical interventions on the spread of COVID-19. ^9–12^ However, the role of such aggregate mobility data in predicting COVID-19 transmission patterns is complex, highly variable over time and space, and notably diminishing since Spring 2020. ^13,14^ Therefore, we conduct extensive data analysis and modeling, to generate novel mobility related variables that explicitly consider trip purpose in addition to broader mobility patterns and incorporate these new mobility-derived metrics into our modeling framework.

For the purposes of this study we obtained aggregated and anonymized mobility data from Safegraph,^15^ a company that provides location data from mobile applications. We generate multiple mobility metrics from the provided weekly patterns and places datasets ^16,17^ as described below.

##### 1.2.1 Mobility Ratio (MR)

We compute a mobility ratio (MR) as a proxy for aggregate mobility movement at population level.^9^ To generate MR, we utilize the following raw point of interest (POI) variables:

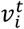: the number of visits to POI *i* on day *t*.

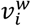: the number of visits to POI *i* during week *w*.

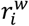: the number of visitors to POI *i* during week *w*.

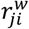: the number of visitors to POI *i* with home location in census block (CBG) *j* during week *w*.

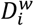: the number of devices residing in given CBG *i* during week *w*. where *t* to represent daily resolution and *w* to represent weekly resolution. The raw data include the number of visits to each POI at daily resolution. However, there is a gap in that the origins of those visits are missing. Hence, additional data preprocessing is needed to estimate origin-destination metrics. For each POI, we first compute the number of visits per visitor 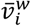 as 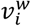 divided by 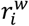, and we assume that 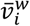 is a constant for all visitors to POI *i* during week *w*. Then we aggregate the visitor’s home location to state-level, and normalize the counts by the state population (*pop*_*c*_) as:

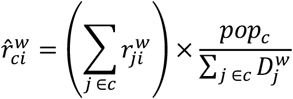

Here 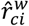 indicates the normalized number of visits from state *c* to POI *i* during week *w*. The probability 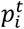 that a visit during week *w* happens during day *t* is calculated as 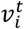 divided by 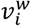, and we assume this distribution holds for visitors from any state. The daily mobility metric 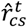 from state c to state s can be estimated as:

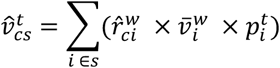

Note that SafeGraph’s data is collected based on device’s home location, so more rigorously, 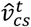 should be interpreted as number of visits with visitors’ home location in state *c* to state *s* on day *t*.

The MR is then defined as:

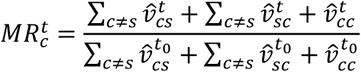

Where 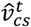 represents the number of trips from location state *c* to state *s* on day *t. t*_0_ represents the baseline time period, which is chosen as the average day of week (e.g., Monday) over the month of February 2020, and 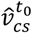 represents the baseline trip rate between locations *c* and *s*.

##### 1.2.2 Importation risk (IR)

In addition to the general mobility trend variable (MR), we generate an importation risk (IR) variable to capture the potential risk of infected visitors arriving at a given destination. This variable combines the real time mobility data and regional case incidence rates at the origin of travel to generate an incidence-weighted travel risk posed to the destination location. The formulation is defined as follow:

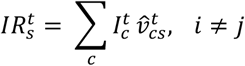

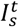 represents the 7-day moving average of reported case incidence rate in trip origin state *i* on day *t*, and 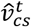 is the same as described above.

##### 1.2.3 Purpose-specific visits (VR)

For each POI, SafeGraph also provides a NAICS (North American industrial classification system) code, which clusters the POIs into different categories based on their primary activity. Previous study^11^ has listed top 50 categories accounting for the largest fraction of visits, we select top 21 as our target destinations, where each type of POI consists at least 1% of overall visits. We generate 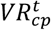 for 21 types of POI (*p* = 21), each one of them is a time series on a daily basis.

For each selected type of POIs (*p*), we estimate the mobility metric 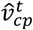 from state c to POI type p as:

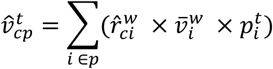

All the selected POI categories are listed below:

**Supplementary Table 1:** The 21 selected POI categories and their NAICS code.

| POI categories | NAICS code |
| --- | --- |
| Full-Service Restaurants | 722511 |
| Limited-Service Restaurants | 722513 |
| Elementary and Secondary School | 611110 |
| Other General Merchandise Store | 452319 |
| Gas Station | 4471 |
| Fitness and Recreational Sports Center | 713940 |
| Grocery Store | 4451 |
| Cafes & Snack Bars | 722514, 722515 |
| Hotels and Motels | 721110 |
| Religious Organizations | 813110 |
| Nature Parks and Other Similar Institutions | 712190 |
| Hardware Store | 444130 |
| Department Store | 452210 |
| Child Day Care Service | 624410 |
| Offices of Physician | 6211 |
| Pharmacies and Drug Store | 446110 |
| Sporting Goods Store | 451110 |
| Automotive Repair and Maintenance | 8111 |
| Used Merchandise Stores | 453310 |
| Colleges, Universities, and Professional Schools | 6113 |
| Convenience Store | 445120 |

An example visualization of all purpose-specific visits metrics for New York State is shown below:

**Supplementary Figure 1:**
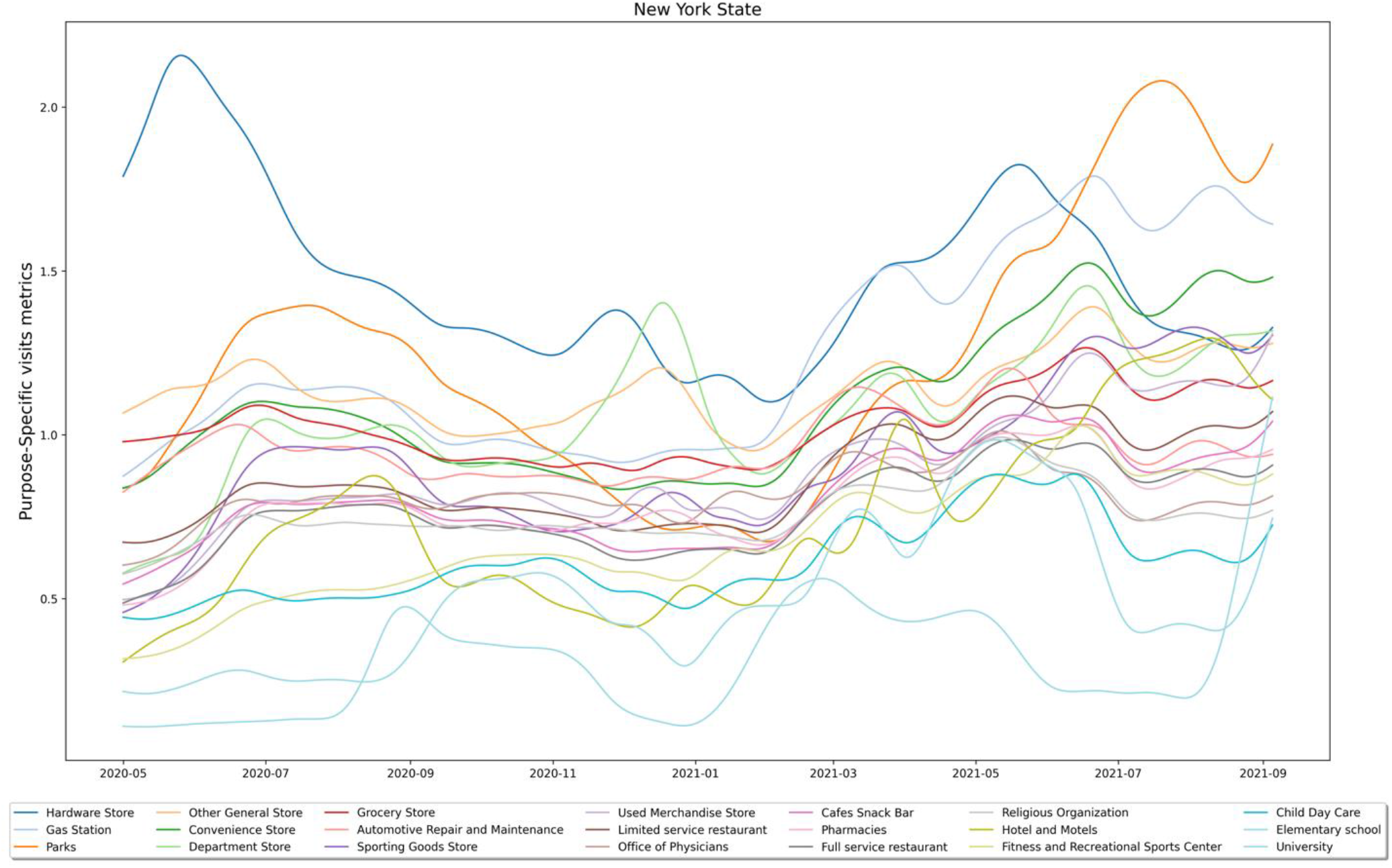
GAM smoothed timeseries of 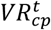 for New York State from May 2020 to September 2021.

Similar to the definition of MR, we define a visit ratio (VR) for each pair of locations (states or counties) and select types of points of interest (POIs). This variable is designed to disaggregate the mobility data by trip purpose, and explicitly considered different travel purposes (work, school, restaurant visits, etc.) within the modeling framework:

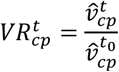

Here, 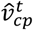 are the estimated daily visits from location (a state or county) *c* to selected POI *p*. Again, *t*_0_ represents the baseline time period. 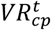 indicates how frequent people visit certain types of destinations relative to the baseline. 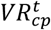 for New York State are shown in Appendix Figure 1, where 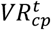 are smoothed with Generalized Additive Model (GAM). The SafeGraph’s data was updated daily during 2020; however, in 2021, the data is updating once a week on every Wednesday.

##### 1.2.4 Principal component analysis of purpose-specific visits metrics

To avoid the highly correlated features and increase computational efficiency, we applied the principal component analysis (PCA)^18^ to all VR variables and select the first five principal components as inputs for the model. By doing this, we could first avoid using similar features that are highly correlated; second, the computational cost is reduced. Here we presented one example of this data preprocessing routine for the first week of September 2021. The average correlations between all the visits metrics across 50 states are show in the Appendix Figure 2 below:

**Supplementary Figure 2:**
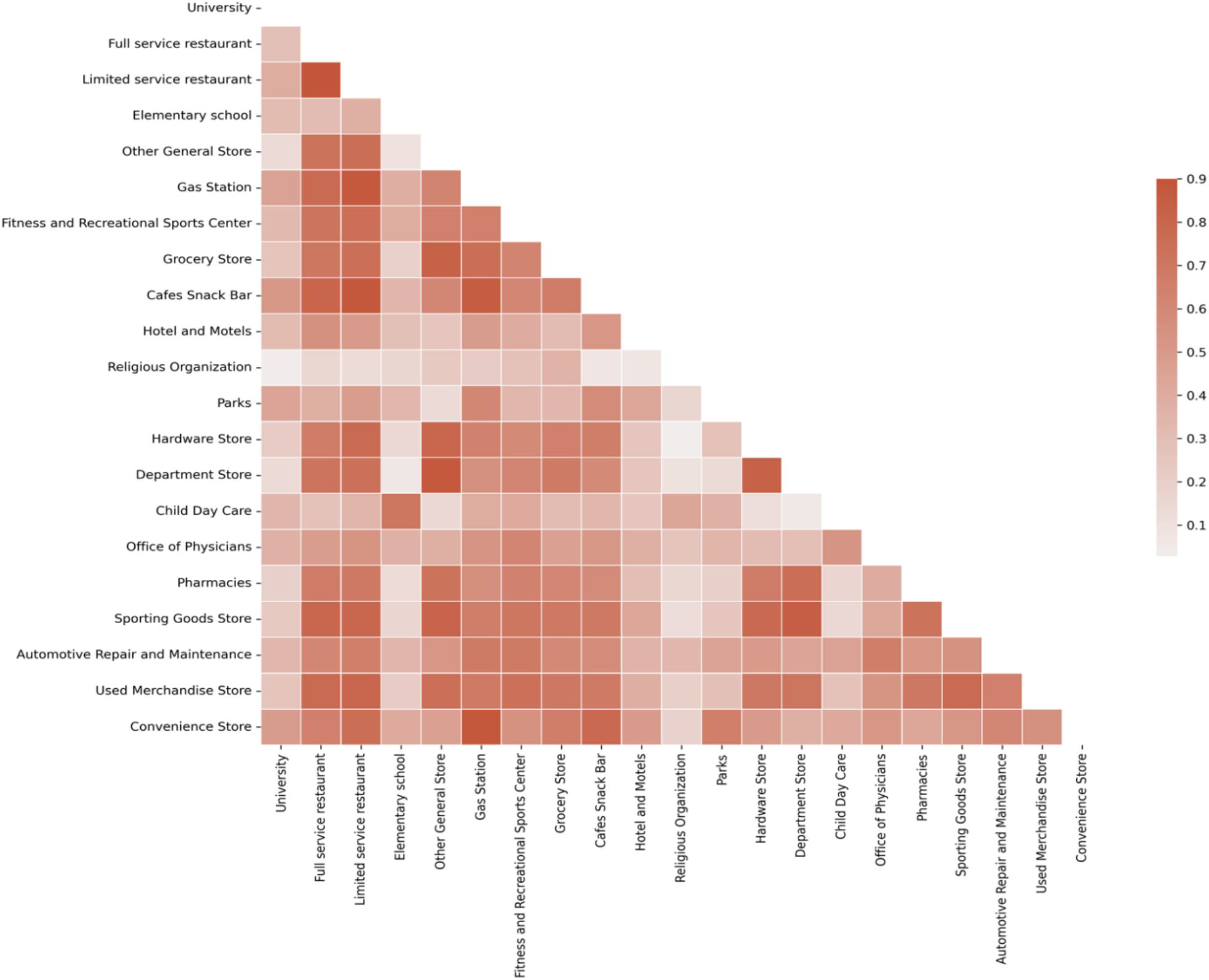
The average correlations between all the visits metrics across 50 states. The color scales represent the magnitude of correlation coefficient, the deeper the color, the higher the correlation coefficient.

All the Pearson correlation coefficients are greater than 0, since all types of visits are more or less affected by lockdown and reopen policy. In addition, there are a few pairs of visits with a correlation above 0·8, which indicates that a feature selection step is necessary.

PCA is a technique to map a higher dimensional data to a lower dimension, and the variance in the lower dimension should explain most of the variations for the full data space. The principal component (PC) is a linear combination of all the features *X*, which can be expressed as:

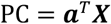

And the covariance matrix **Σ** of PC is:

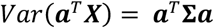

The goal of PCA is to preserve the original variance as much as possible, hence *Var*(***a***^*T*^***X***) should be maximized under the condition that ***a***^*T*^***a*** = **1**. Using the Lagrange’s multiplier, we could generate each PC as follow:

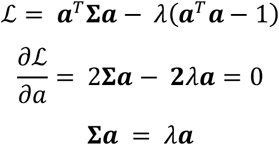

Hence ***λ*** and ***a*** are the eigenvalues and eigenvectors of covariance matrix **Σ**. Therefore, consider a dataset with *p* variables, the *i*th principal component and its variance are:

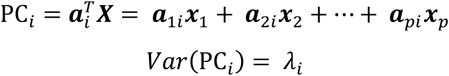

Here ***a***_*i*_ are the desired weights assign to each feature. The total amount of variance explained by first *i* components is:

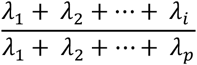

The variance explained by the first five PCs are each state is shown in Appendix Figure 3 below, where x-axis is the percentage of variance explained and the red dashed line represents 80%.

**Supplementary Figure 3:**
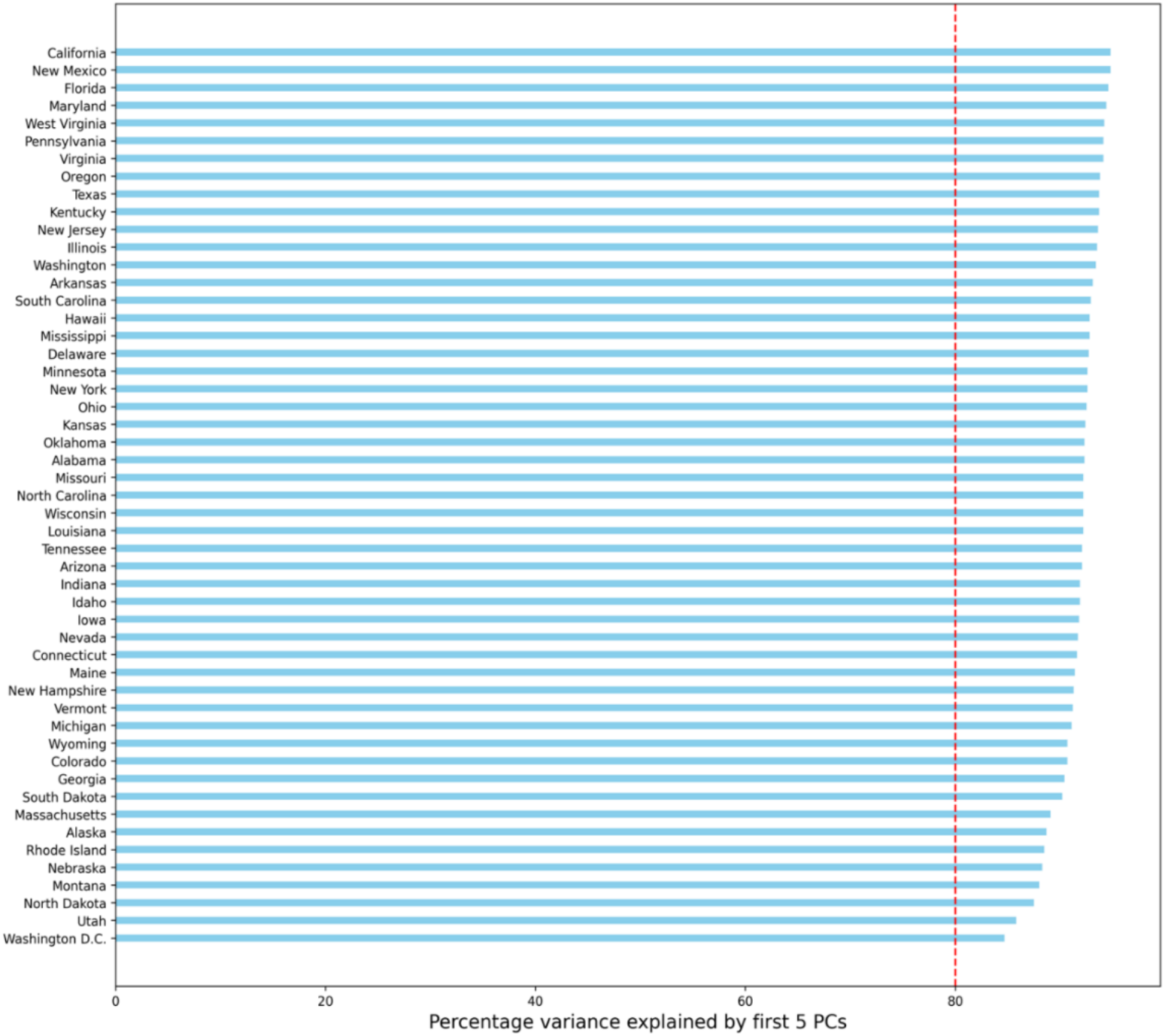
Percentage variance explained by the first 5 PCs for all the states. The red dot vertical line represents 80% variance explained.

#### 1.3 COVID-19 symptoms survey data

Human behavior, while evasive to quantify, is thought to play a dominant role in the outbreak patterns observed for COVID-19. Various public survey efforts exist to generate behavioral indicators that can be used to better infer transmission dynamics, and these have been illustrated to improve predictive accuracy compared to those without them.^19^ In this work we utilize data from the COVID-19 symptoms survey. The survey is conducted through Facebook’s platform in collaboration with the Delphi group.^20^ From this survey we use “the estimated percentage of people who know someone in their community having Covid-like symptoms”. The timeseries data is provided at a daily resolution and smoothed using a 7-day moving averages to generate our input variable.

#### 1.4 Climate data

Some evidence points to climate and seasonality as potential factors associated with COVID-19 transmission,^21,22^ although its role remains unclear.^23^ To account for the possible impact of climate and seasonality on transmission risk, we include daily population-weighted hydrometeorological data, sourced from the JHU COVID-19_Unified-Dataset GitHub repository.^24^ The full set of variables we consider are near-surface air temperature (°C), and the total precipitation (mm/day).

#### 1.5 Demographic data

COVID-19 is known to have a disproportionate impact across demographic groups, specifically age and race.^25^ For this reason, we included total population and population percentage over 65 years of age as two separate static variables in our model. The population data were collected from the American Community Survey of the US Census Bureau.^26^ We use the 2019 Single Year of Age and Sex Population Estimates dataset to calculate the percentage of the population over 65 years old for each state.

#### 1.6 SARS-CoV-2 Genomic Surveillance data

Of particular interest in this study is the potential value of SARS-CoV-2 genomic surveillance data in forecasting models to predict surges that may be driven by new variants in a more accurate and timely manner, or more generally, the impact new variants might have on COVID-19 transmission patterns. To address this research questions, we conduct a case study that utilizes COVID-19 genomic data downloaded directly from GISAID.^27^ From the available set of sequences, we calculate the proportion of each variant (theoretically in circulation) each day over the course of the pandemic.

Based on the sample collection date (October 7th, 2021), we select the United States data at the state level between April 1, 2021, to end of August 2021. First, we calculated the coverage rate as the total number of genomic samples over the number of confirmed cases for each state during the selected period. Then, we filtered out the states with less than 5% of coverage. We selected all the State with at least 5% of overall sampling coverage. As shown in Appendix Figure 4, all the states with color have sampling coverage higher than 5%, while the states in grey have coverage less than 5%.

**Supplementary Figure 4:**
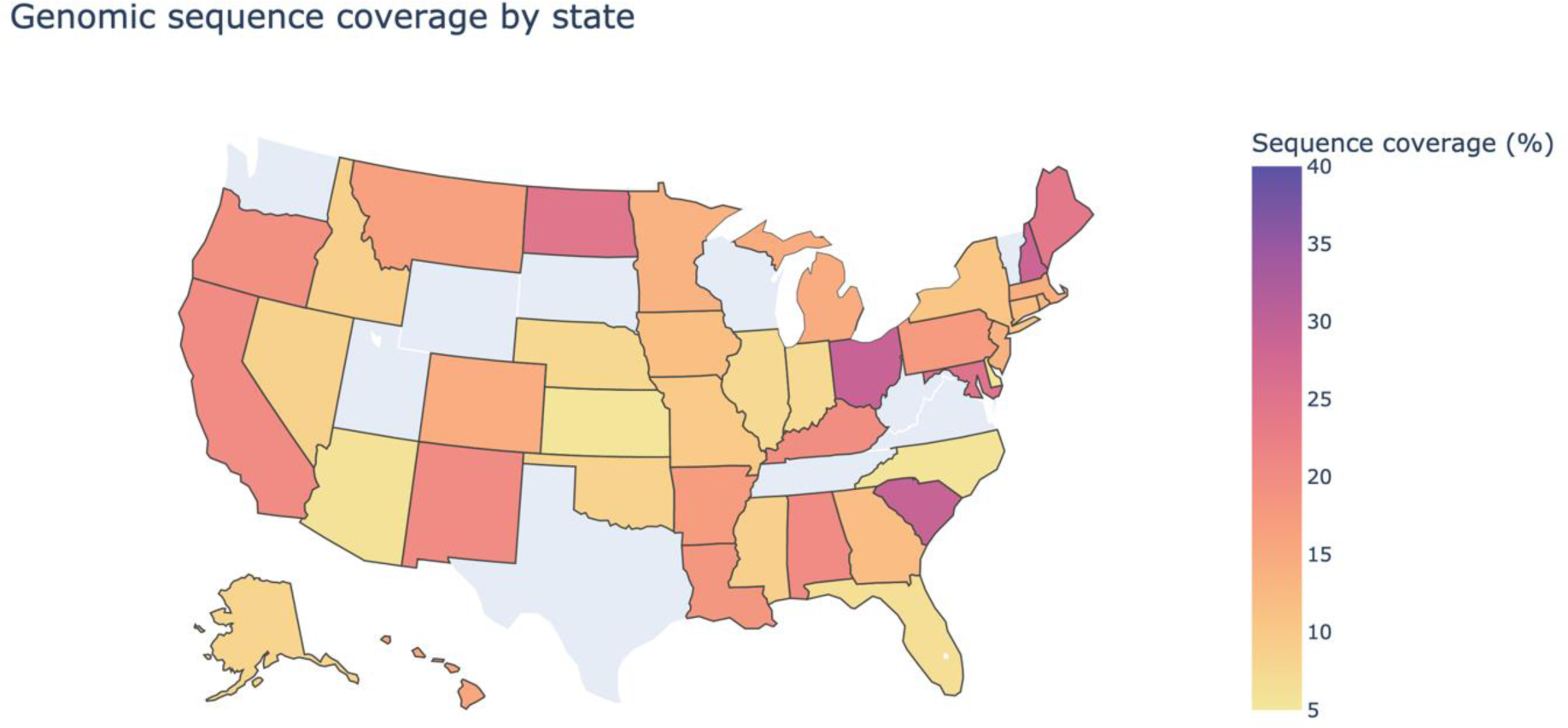
Genomic sequence coverage by state between April 1st, 2021 and August 21st, 2021. The color scales represent the magnitude of the sequence coverage, the deeper the color, the higher the coverage.

We generate time series of lineage proportion for the Delta, Gamma, and Alpha variants among all the virus lineages. Then, all the rest of the lineages are classified as others. The four proportion time series are smoothed with a 7-day moving average. We assume that this proportion of variants samples also apply to the proportion of variant cases within the confirmed cases. The genomic features are defined as:

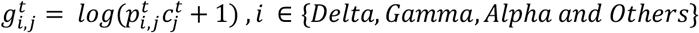

Where 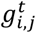 is estimated logarithm of confirmed cases for variant group *i* for state *j* at time *t*, 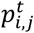 is the proportion of variant group *i* for state *j* at time *t* and 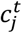 is the confirmed cases for state *j* at time *t*.

We calculate the collection to submission time for all collected genomic data by October 7th, 2021. The distribution of CST is shown in Appendix Figure 5, with a median CST of 24 days. In our case study, we apply a scenario analysis to test the value of the genomic data, we assume all the genomic data will be available with a CST of 7 days. By doing this, we want to provide a proof of concept that the timely genomic data (7-days lag) is valuable for short-time COVID-19 forecasting.

**Supplementary Figure 5:**
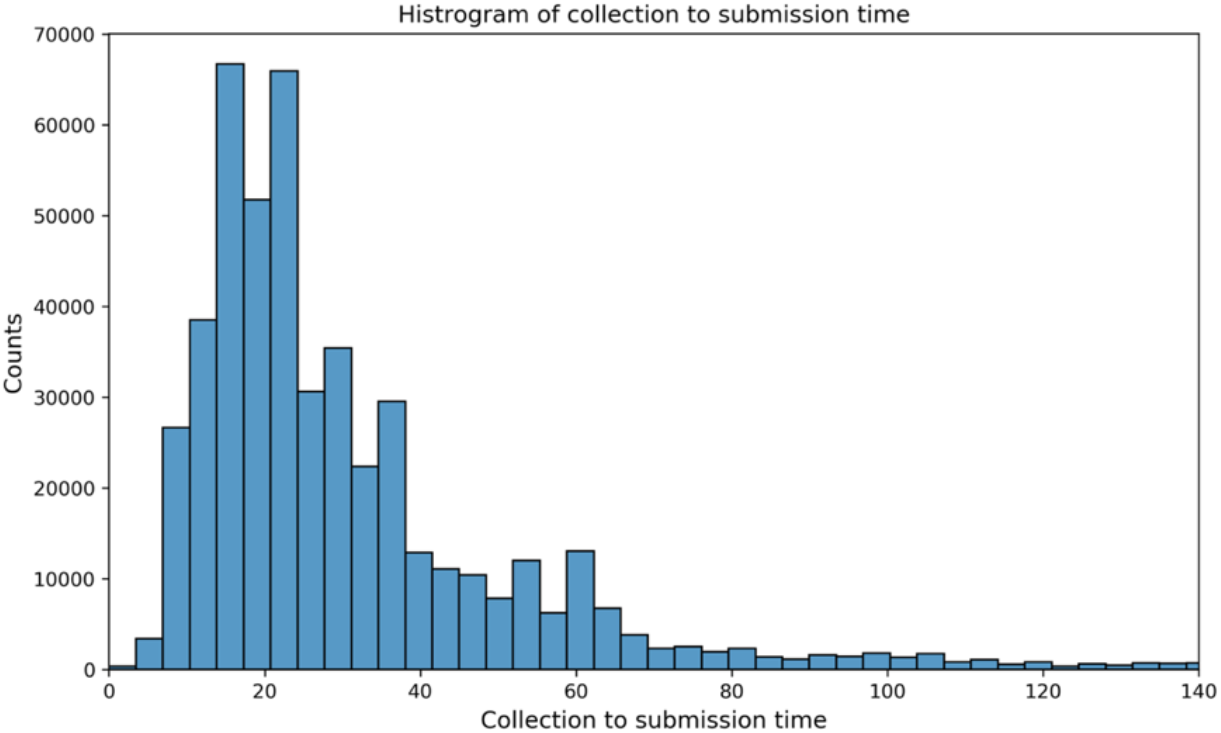
Histogram of collection to submission time as October 7th, 2021

The proportion of selected variant is shown in Supplementary Figure 6. Each dot represents the raw proportion on a given day and each solid line represents the trend smoothed by Generalized Additive Model (GAM). The deeper the color, the earlier the Delta variant became dominant in that state. The timeseries illustrate the quick rise of the Delta variant from May 15 and July 15, at which time it because the dominant lineage across the U.S., soon converging to 100%. However, by the end of August, the Delta variant was already being replaced by the Omicron variant (which occurred outside this study period).

**Supplementary Figure 6:**
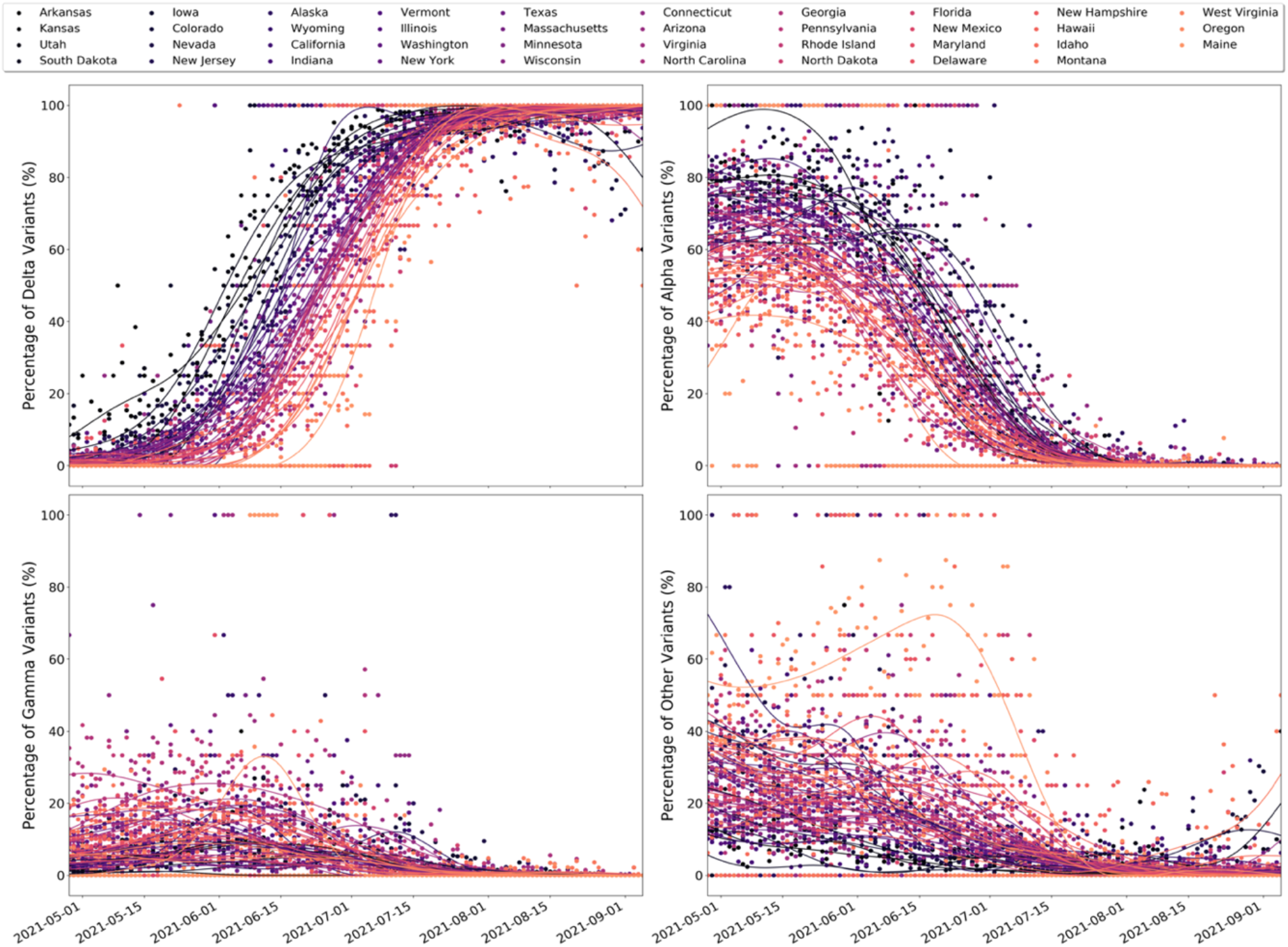
Time varying proportion of selected variant for 39 states in the U.S. Colors represent data for different state. The dots represent the raw genomic data while the plotted lines are smoothed with a GAM.

### 2. Supplementary method

#### 2.1 An example implementation of the multi-stage LSTM model

The multi-stage frame train the framework to predict reported cases/deaths and other time series input for the next 7 days. At the initial stage, the model uses the most recent data as input, then at the later stage, the model adapts previous prediction as input to make further predictions. For example, if we wish to predict the number of new COVID-19 cases each week for the next 2 weeks using as input three time-varying features from the weeks prior: incident cases, cases growth rate, and MR, the model stages are set up as follows:

Stage 1: We use observed data from the last three weeks, specifically day (*t* − 21) to day *t* as the length of the sequence of the input data for both the main and feature model. The outputs from the main model are incident cases in the week following, *t* + 1 to *t* + 7 (total predicted cases for one-week ahead). The outputs from the feature model are the case growth rate and MR for the same one-week ahead window, *t* + 1 to *t* + 7. Together, the forecasted values from the main and feature model provide the required input for the following stage (stage 2), which aims to predict incident cases for two weeks ahead.

Stage 2: The stage 2 outputs from the main model are incident cases from *t* + 8 to *t* + 14, which are then converted to our second week predicted cases, while the outputs from the feature model are cases growth rate and MR from *t* + 8 to *t* + 14. To generate these 2-week ahead outputs, we require a timeseries from *t* − 14 (observed) to *t* + 7 (unknown at time t), again, as the model requires a three-week length of the sequence of the input data. The stage one output enables the extension of each of the time series from the current time t to *t* + 7. We use a combination of the observed data (*t* − 14 to day t) and outputs from stage 1 (time t to *t* + 7) to generate the required timeseries as input for stage 2. This same process is repeated to generate inputs for each following stage. For any combination of target variables, both the main and feature models are trained on the same dataset and applied simultaneously to generate predictions.

#### 2.2 Formulations of LSTM model

Both the main and feature models introduced above have the same network structure, which is built by one layer of long-short term memory (LSTM) network ^28^ and two layers of Multilayer perceptron (MLP). LSTM is a special kind of recurrent neural network, which is capable of learning long-term dependencies. The key idea behind LSTM is the cell state (*C*_*t*_) controlled by three gates named input gate (*i*_*t*_), output gate (*o*_*t*_) and forget gate (*f*_*t*_). The cell state keeps the information that passes along the sequence, and those three gates help filter information in the cell state at each time point. The output *h*_*t*_ from the LSTM layer will serve as input to pass through MLP layers. Then the finial predictions are the outputs from MLP layers. We apply the dropouts at the MLP layers to include randomness for the predictions.

The LSTM layer can be formalized as follow:

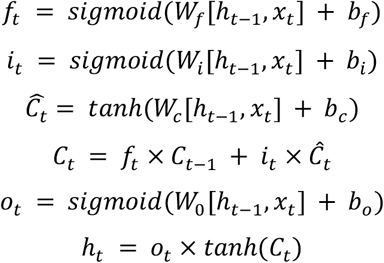

where *x*_*t*_ and *h*_*t*_ denote, respectively, input data and hidden state at time *t. Ĉ*_*t*_ denotes a candidate new cell state that could be added to the cell state. *W*_*j*_ and *b*_*j*_ represent weights and bias terms at each gate or cell state *j*. The prediction function for the entire network model can be formulated as:

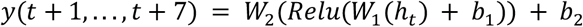

where *y*(*t* + 1, …, *t* + 7) are the predicted cases from *t* + 1 to *t* + 7, *W*_1_, *W*_2_, *b*_1_, and *b*_2_ are weights and biases for the fully connected neural network, *Relu* is the activation function:

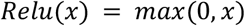

#### 2.3 Model parameterization

The models are implemented using Python 3·8 with open sources packages such as PyTorch, Pandas and NumPy. In initial setting of the multi-stage Neural Network model, different size of hidden layers in LSTM were explored for model training and testing. The sensitivity analysis showed subtle differences of performance between different settings. Hence, we set size of hidden layers as constant for both main model and features model. The full dataset is randomly divided into two sets, 70% for training and 30% for testing, early stop will apply, if the testing error no longer improving. We use smoothed L1 loss as the model loss function, the formulation is described below:

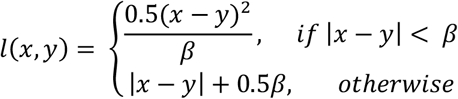

The model performances are most sensitive to the training periods; we test different combinations of training periods for the main model and the features model. Finally, we use 1 layer of LSTM layer connected with two dense connected layers. For the main model, we set the hidden layer size as 256, and for the features model, we set the hidden layer size as 328.

#### 2.4 Model evaluation metrics

The formulations for AE, PAE, and WIS ^29^ are defined below. *F* is the model prediction function; *u, m* and *l* are the upper bound, the median, and the lower bound of the predictions, respectively, and *y* is the ground truth.

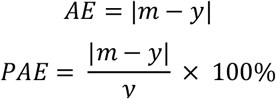

For each (1 − *α*) × 100% prediction interval, the WIS is defined as:

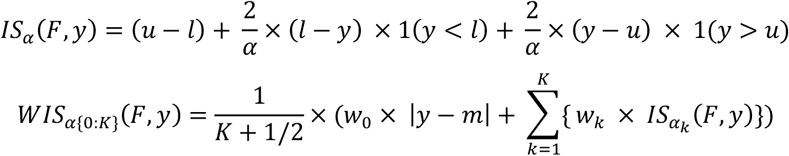

where 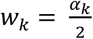, in this paper we choose *K* = 3 with *α*_1_ = 0 · 05, *α*_2_ = 0 · 2 and *α*_3_ = 0 · 5.

#### 2.5 Model Selection

All the results in the main document are based on our best performance model. We apply sensitivity analysis for four alternative models under two time period separately. From August 2020 to Feb 2021, the epidemiological data do not contain vaccination data, while since February 2021 to August 2021, we add the vaccination data to the epidemiological data category. Static variables are automatically included for all the models. The category assignment for each variable can be found in main document table 1.

There are two steps for the model selection: 1) Determine the optimal training periods for each candidate model. 2) Under the optimal training periods, find the best performing model. For this part of the analysis, we evaluated our models based on point predictions.

For each of the four models, we test different training periods for the main model (*t*_*m*_) and the features model (*t*_*f*_). Each prediction is evaluated by PAE and AE. In this study, we weighted them equally in model selection. The rule for selecting the best training period is described below:

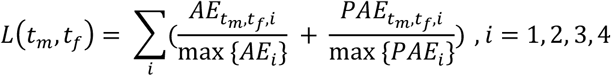

Where *L*(*t*_*m*_, *t*_*f*_) is an aggregated loss function of hyperparameters *t*_*m*_ and *t*_*f*_; 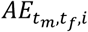 and 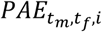 are average absolute error and average percentage absolute error at forecasting window *i*; max {*AE*_*i*_} and max {*PAE*_*i*_} are maximum errors corresponding to all the combinations of (*t*_*m*_, *t*_*f*_). We select *t*_*m*_ and *t*_*f*_ that minimize the *L*(*t*_*m*_, *t*_*f*_) (Supplementary Table 2).

**Supplementary Table 2:** Summary of training periods selection.

| Input features | Time Period | $t_m$ | $t_f$ |
| --- | --- | --- | --- |
| Epidemiological data | August 2020 to February 2021 | 60 | 45 |
| Epidemiological data | February 2021 to August 2021 | 30 | 30 |
| Epidemiological, and mobility data | August 2020 to February 2021 | 60 | 75 |
| Epidemiological, and mobility data | February 2021 to August 2021 | 40 | 40 |
| Epidemiological, mobility, and survey data | August 2020 to February 2021 | 45 | 75 |
| Epidemiological, mobility, and survey data | February 2021 to August 2021 | 30 | 30 |
| Epidemiological, mobility, survey, and climate data | August 2020 to February 2021 | 60 | 45 |
| Epidemiological, mobility, survey, and climate data | February 2021 to August 2021 | 40 | 40 |

Once the training periods were determined, we further evaluated the performances between different inputs. The results of comparing these four models with the CDC ensemble model from August 2020 to August 2021 are shown in Appendix Figure 7 and 8 reveal the break down performance before and after February 2021. Both PAE and AE are used to evaluated performance, and the value for each bar in the plots is the average over all states and time.

**Supplementary Figure 7:**
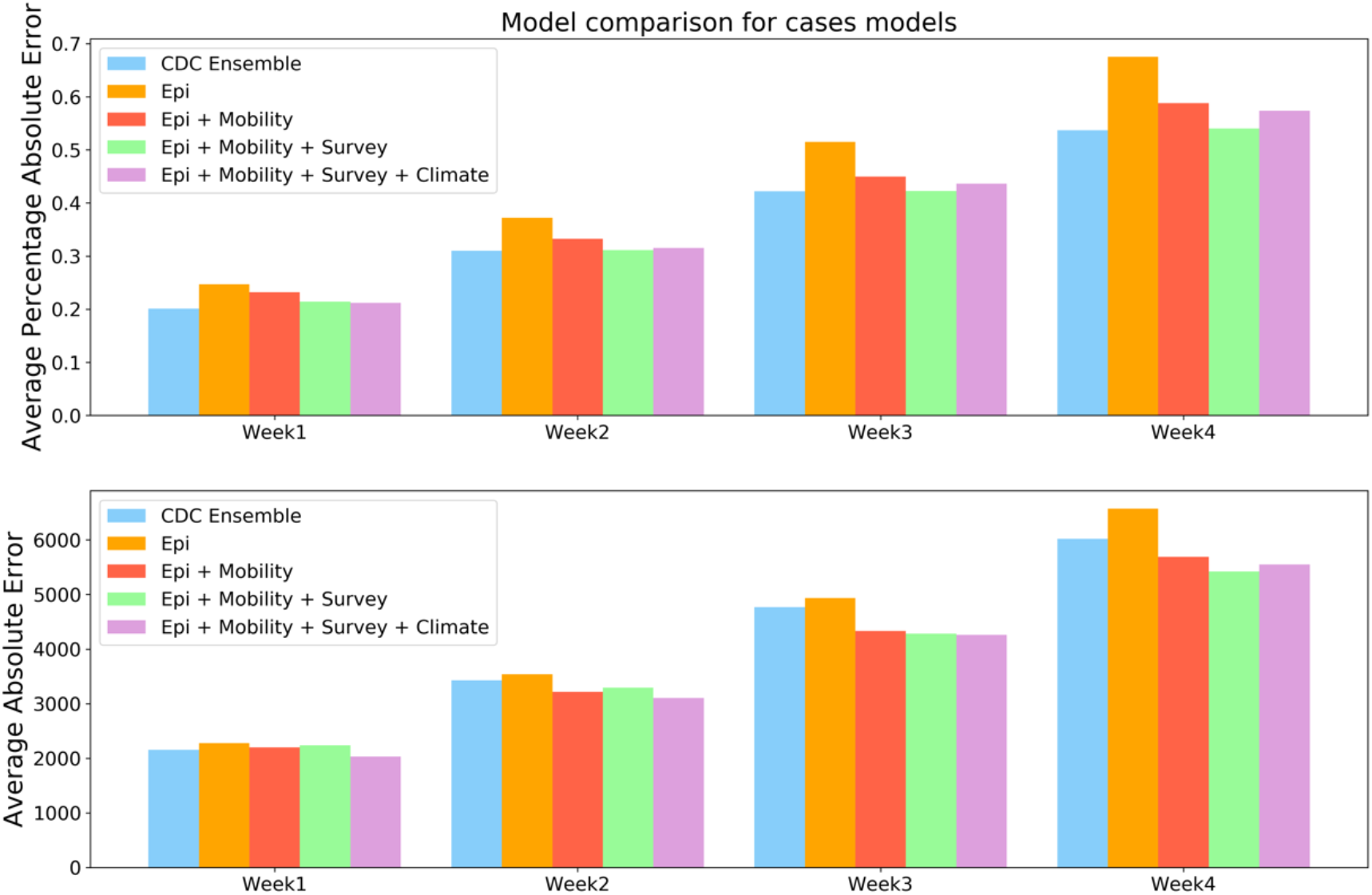
Model comparison for cases models by the mean PAE and the mean AE for the period between August 2020 to September 2021. The y-axis represents the average absolute error for 1-4 weeks cases’ prediction results across the entire study period.

**Supplementary Figure 8:**
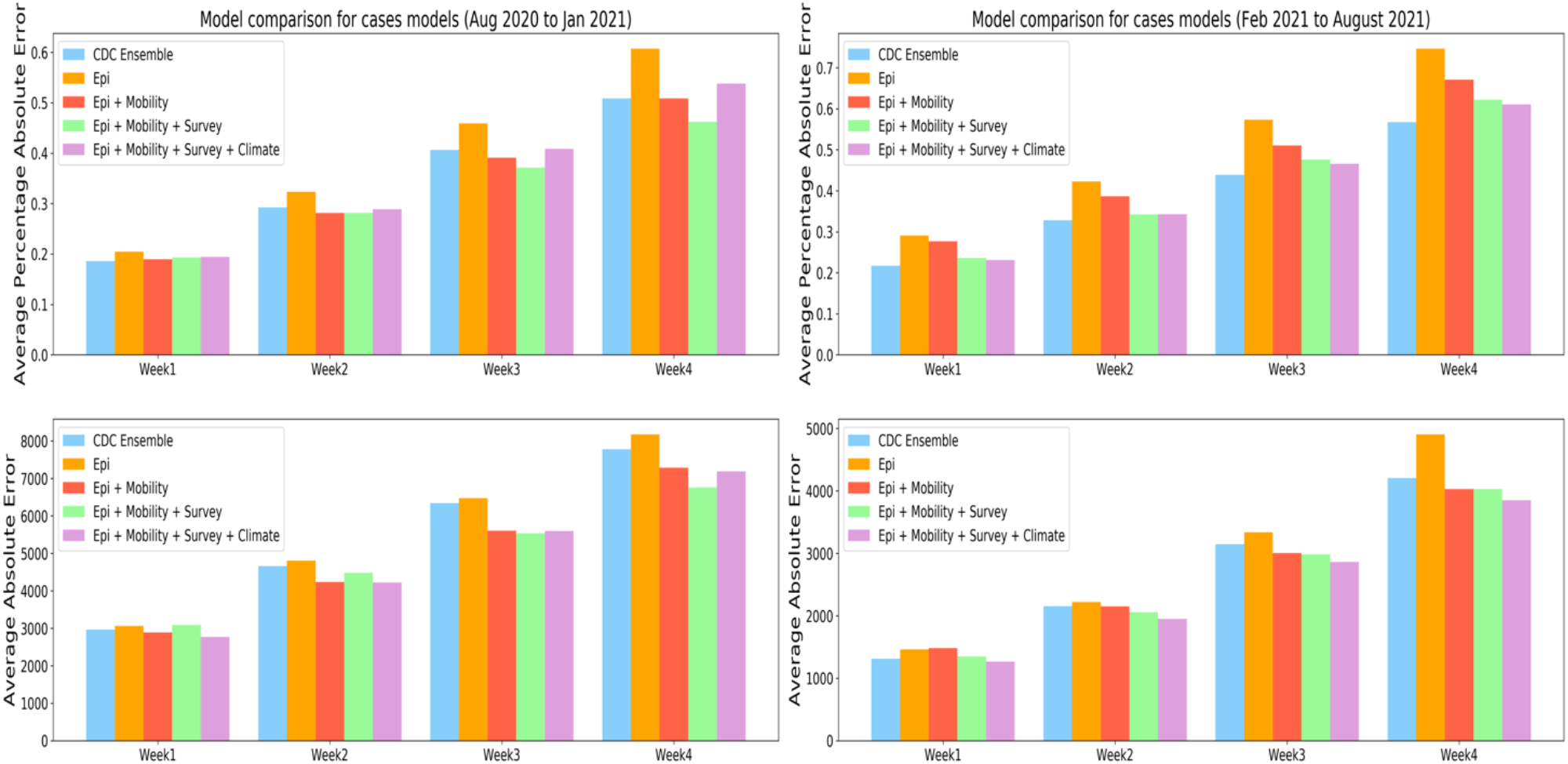
Model comparison for cases models by the mean PAE and the mean AE during two periods: from August 2020 to February 2021, and from February 2021 to August 2021. The y-axis represents the average absolute error for 1-4 weeks cases’ prediction results across selected period.

### 3. Supplementary results

#### 3.1 Model performance across time by AE and WIS

Appendix Figure 9, and 10 illustrate the relative performance of the LSTM against the CDC ensemble model for each of the 52-week periods evaluated, for 1 to 4 week forecast windows, based on AE and WIS respectively. Each pair of bar plots represents PAE distribution for all the states at a given week, where the green bar represents the error distribution for the multi-stage LSTM model, and the yellow bar represents the error distribution for the CDC ensemble model. The red curve represents the weekly reported cases at the national level.

**Supplementary Figure 9:**
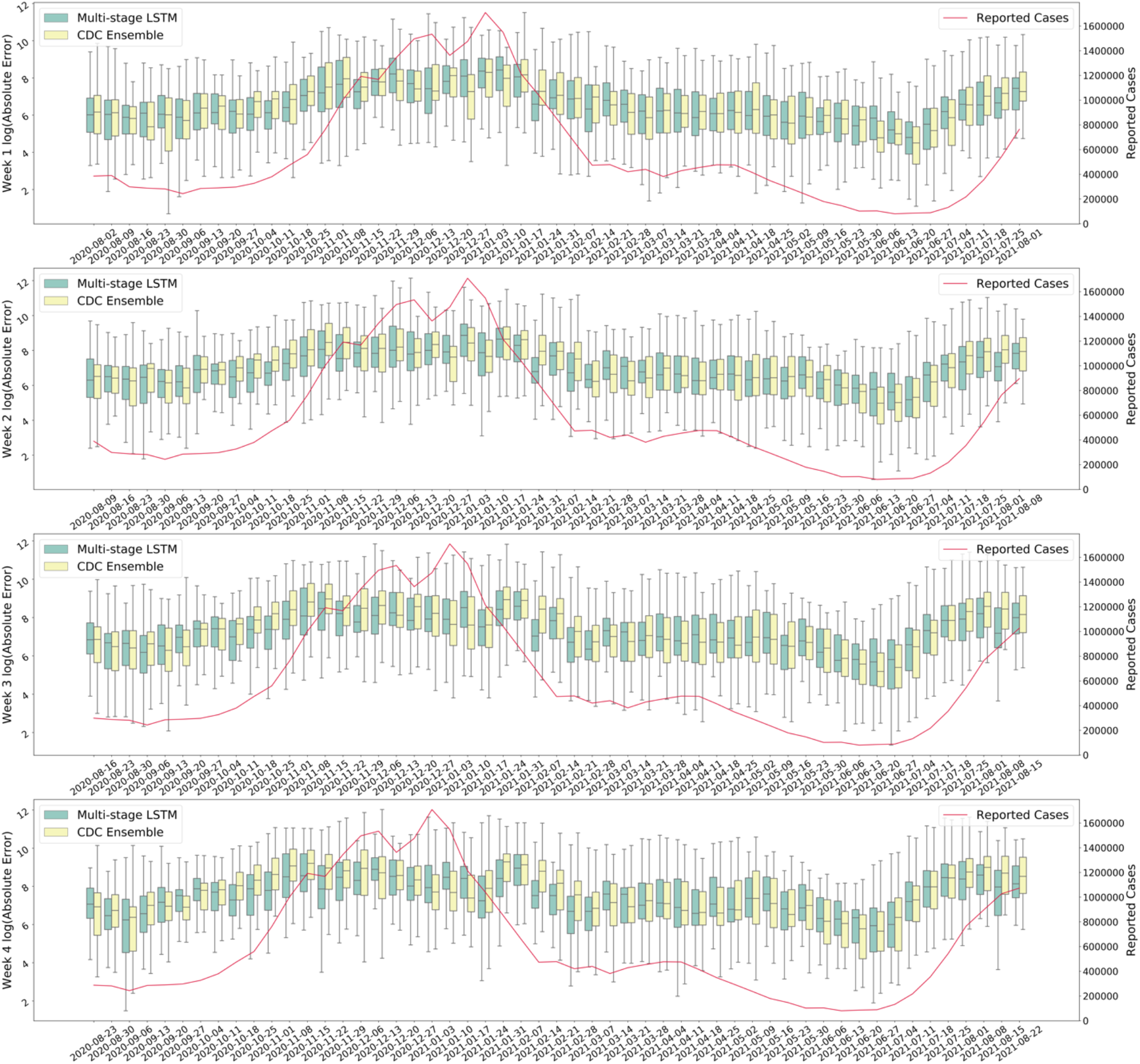
Comparison of model performance (AE) between the multi-stage LSTM Model and the CDC ensemble model. For better visualize the results, we normalize AE by taking the log. The left y-axis represents the log AE for 1-4 weeks cases’ prediction results and right y-axis represents national level reported cases.

**Supplementary Figure 10:**
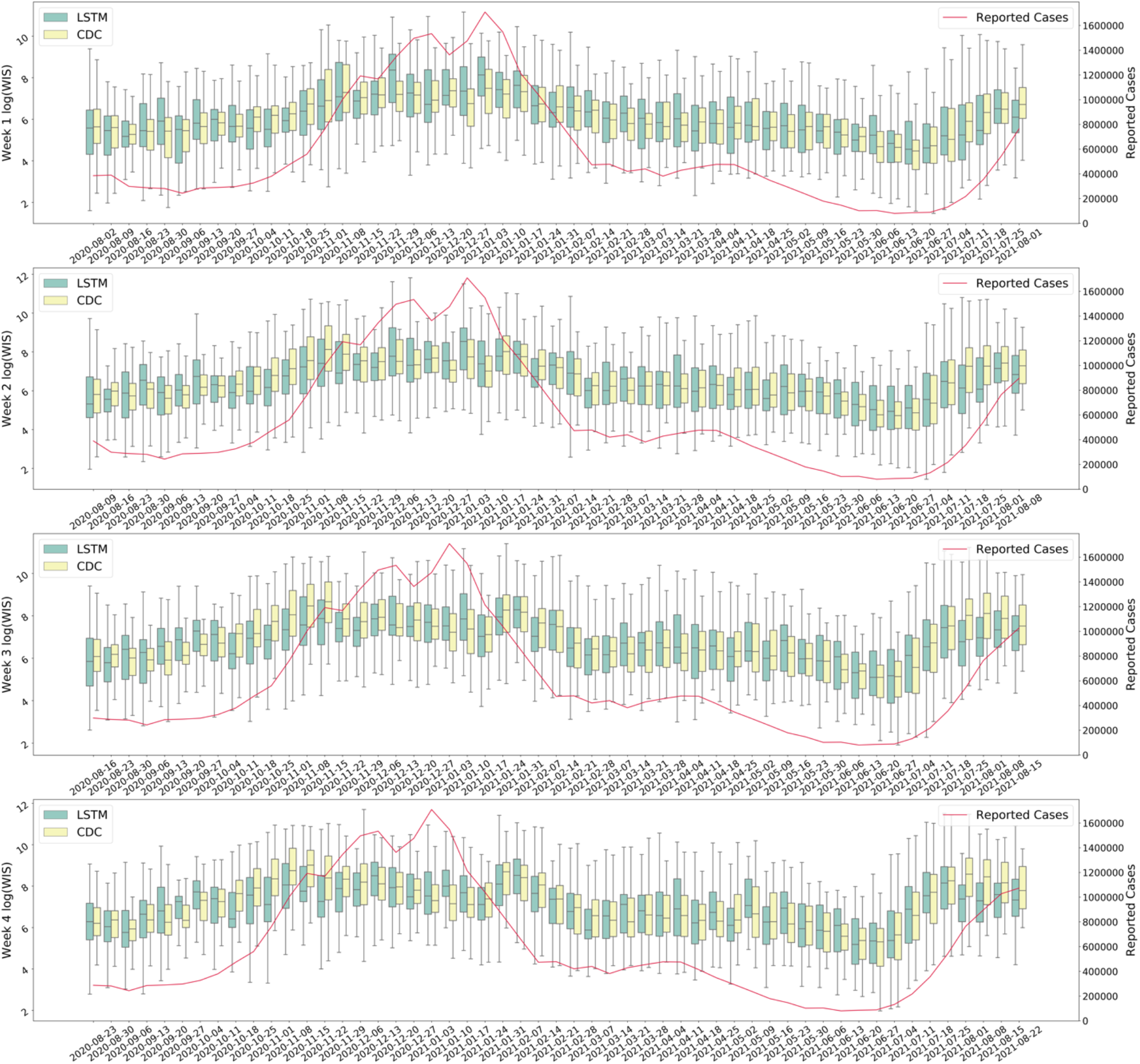
Comparison of model performance (WIS) between the multi-stage LSTM Model and the CDC ensemble model. For better visualize the results, we normalize WIS by taking the log. The y-axis represents the log WIS for 1-4 weeks deaths’ prediction results.

#### 3.2 Model performance across states by AE and WIS

The Pearson’s correlation coefficient for raw WIS and the state-level population is 0·96. Thus, we normalized the WIS by population to remove population bias. For visualization purpose we convert AE into logarithm scale. The color scales represent the magnitude of each error metric; the scales are fixed as (1,10) and (6,11) for WIS and normalized log(AE), respectively. Hence, the deeper the color, the larger the error for the state.

**Supplementary Figure 11:**
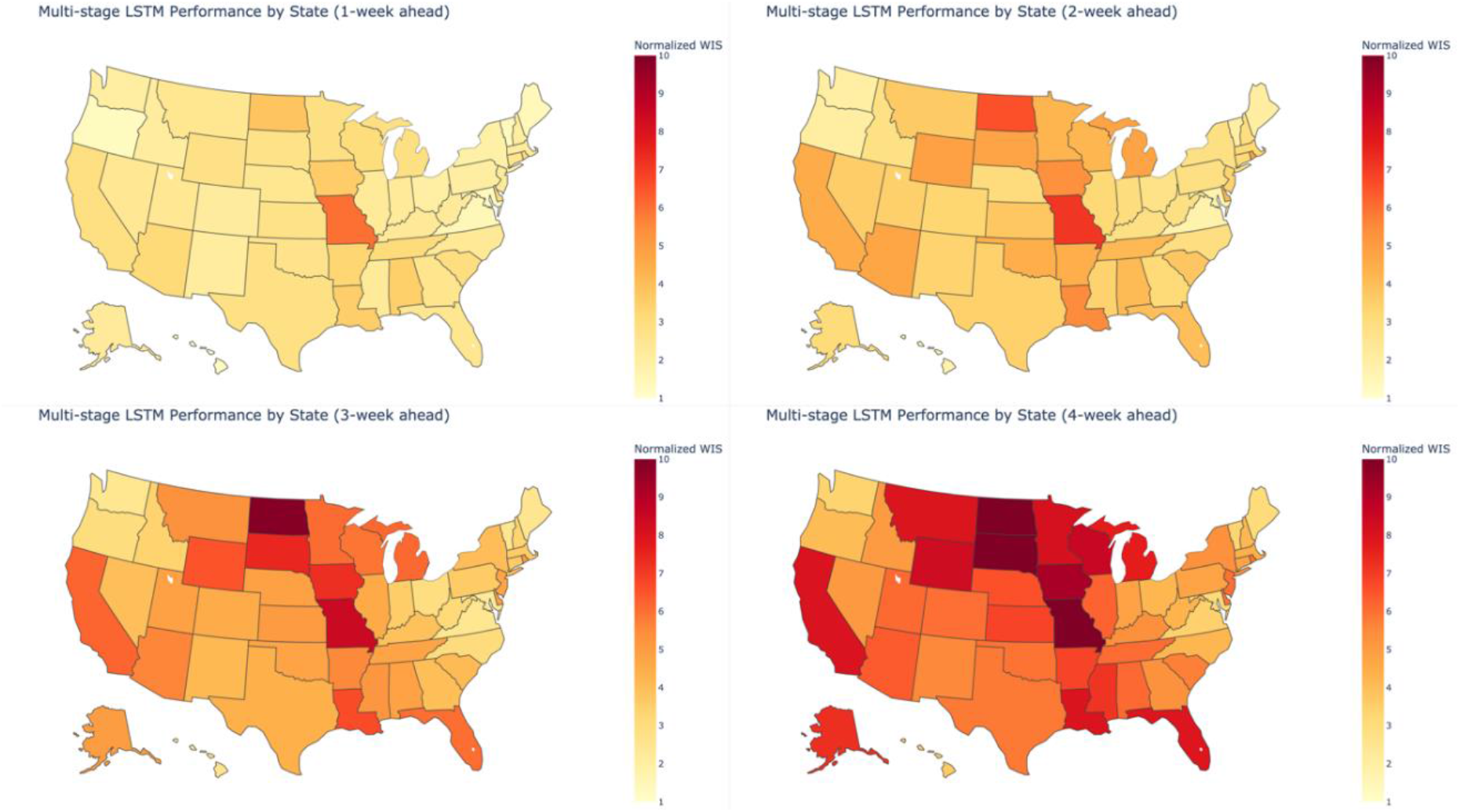
Average model performance at the state level by normalized WIS. The color scales represent the magnitude of the error metric; the scales of normalized WIS are fixed in 1–10 range. The deeper color corresponds to larger error.

**Supplementary Figure 12:**
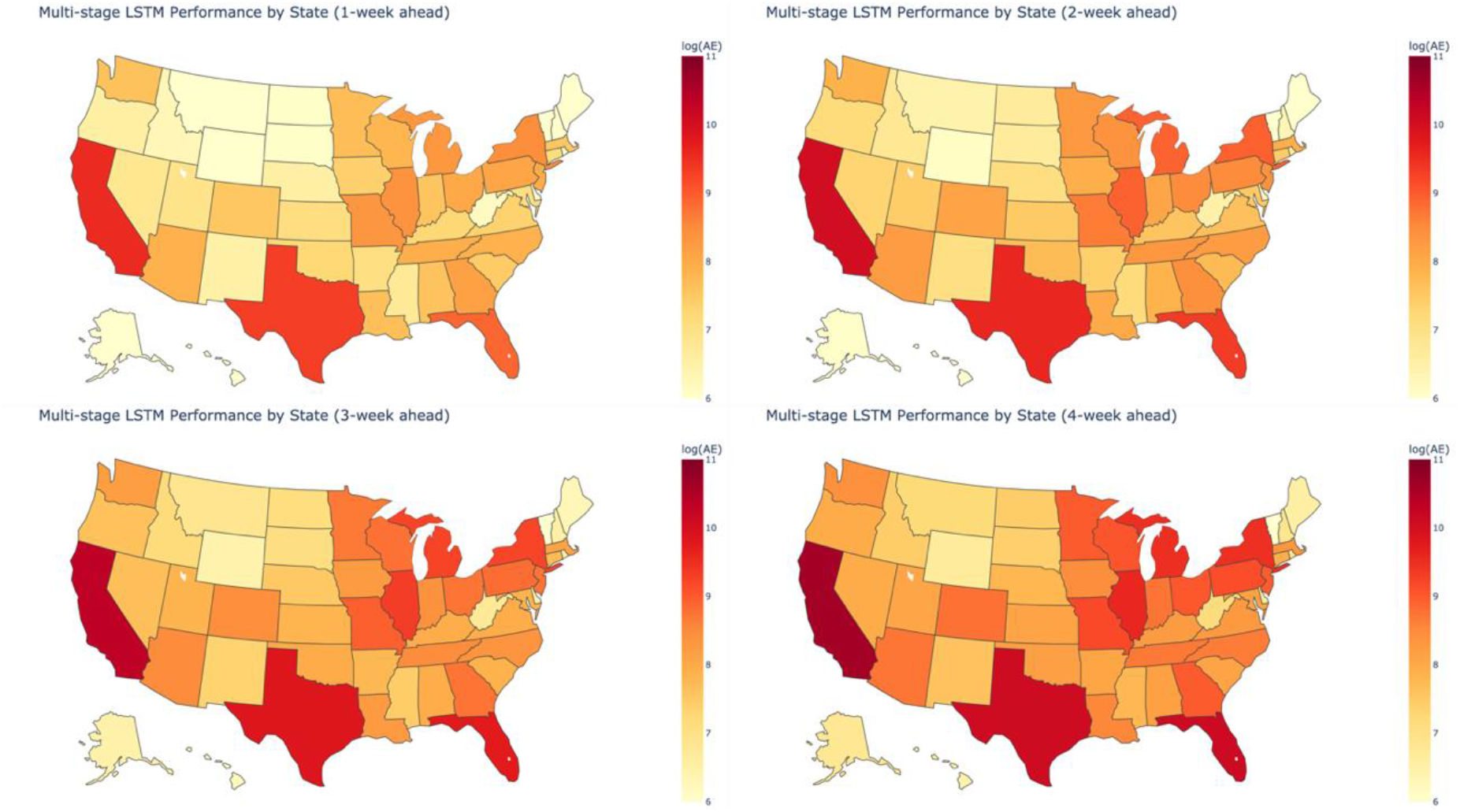
Average model performance at the state level by logarithm of AE. The color scales represent the magnitude of the error metric; the scales of logarithm AE are fixed in 6–11 range. The deeper color corresponds to larger error.

#### 3.3 Reported Cases Trend by Region

To better understand the presence of spatial patterns in model performance as illustrated in Figures 3, we generate and compare state-level confirmed case trends between each region.

We first group states into each HHS region as shown in Appendix Figure 13 below. Most of the midwestern states are included in region 5, region 7, and region 8.

**Supplementary Figure 13:**
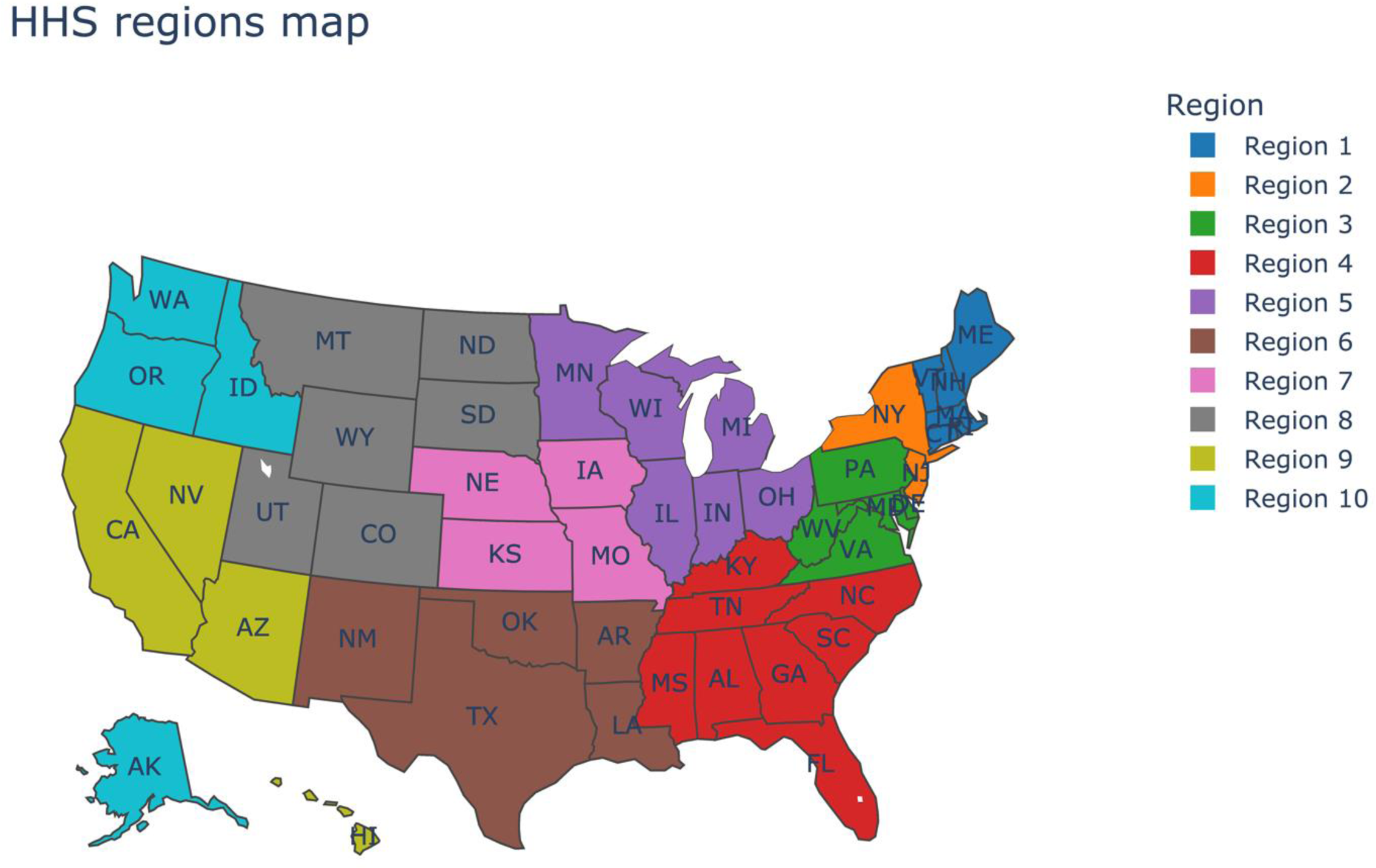
HHS regions map.

We define a trend variable 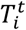 for each state *i* at each week *t* using the weekly confirmed cases from August 2020 to August 2021 for each state as follows:

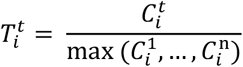

where 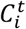 is the number of confirmed cases for state *i* at week *t*, and max 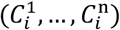 is the maximum weekly confirmed cases over the selected weeks 1 to *n*, for state *i*. 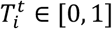 represents how reported cases for state *i* at week *t* compare to the highest reported cases over the entire period.

For each HHS region *j*, we calculate the average trendline 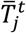 based on all state trends within the region:

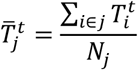

where *N*_*j*_ is the number of states within HHS region*j*. This variable represents the normalized reported cases trend for each HHS region (i.e. when the most of states within a region show decreasing or increasing trend). The normalized 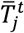 variable enables better comparison of trends across regions since 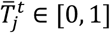.The results of 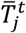 are shown in Appendix Figure 14.

**Supplementary Figure 14:**
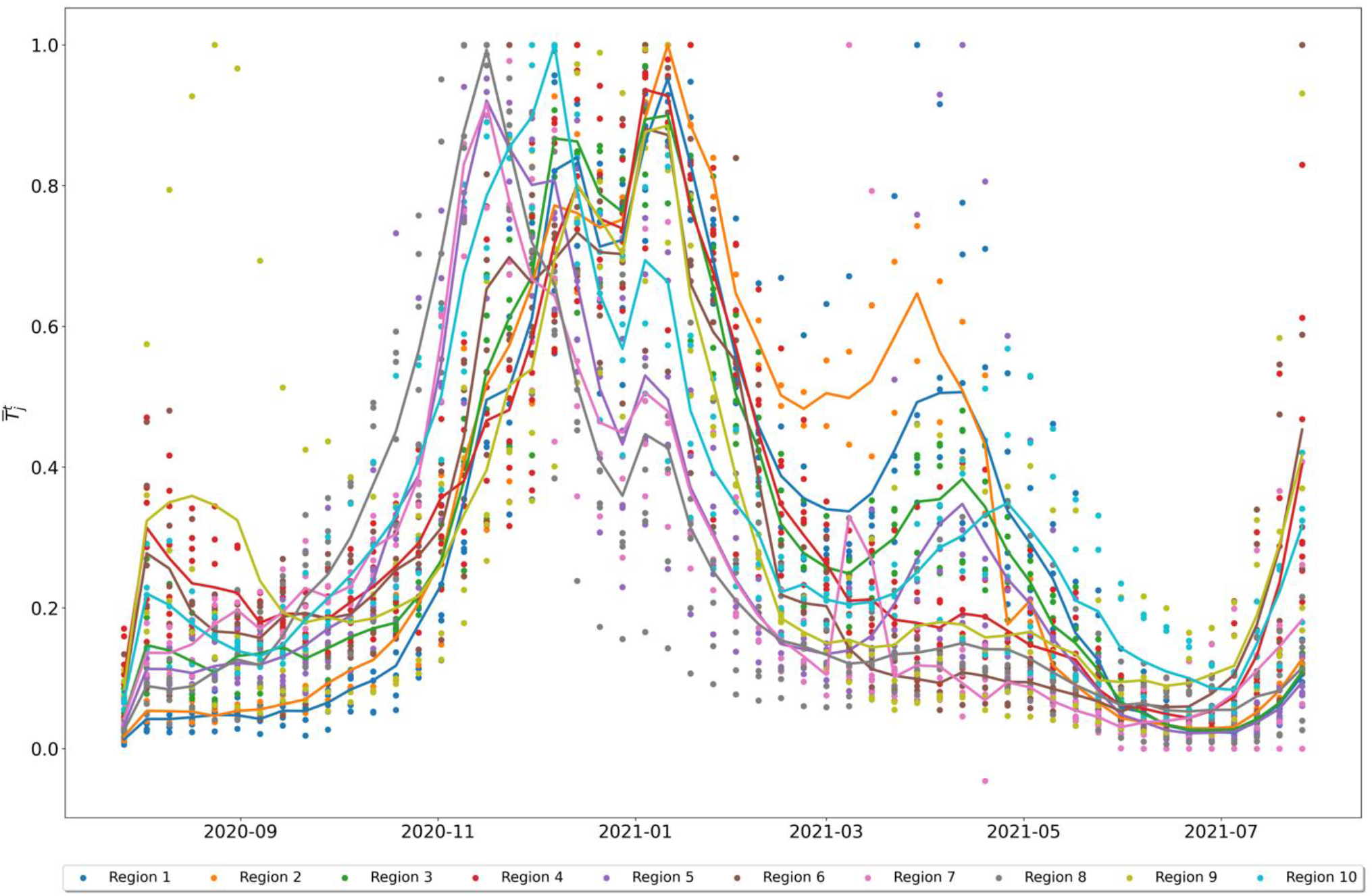
Weekly confirmed cases trend 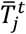 by HHS regions.

Most of the midwestern states are included in region 5, region 7, and region 8. This plot illustrates different transmission patterns for regions 5, 7 and 8, relative to the rest of the country between November 2020 and February 2021. Specifically, while most of regions have upward 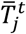, the three regions mentioned above have decreasing trends. This result could be a possible explanation for why the forecasts for the midwestern states is less accurate relative to the rest of the country (as shown in Figure 3 and Appendix Figure 11).

#### 3.4 An example of classification for different outbreak phases

**Supplementary Figure 15:**
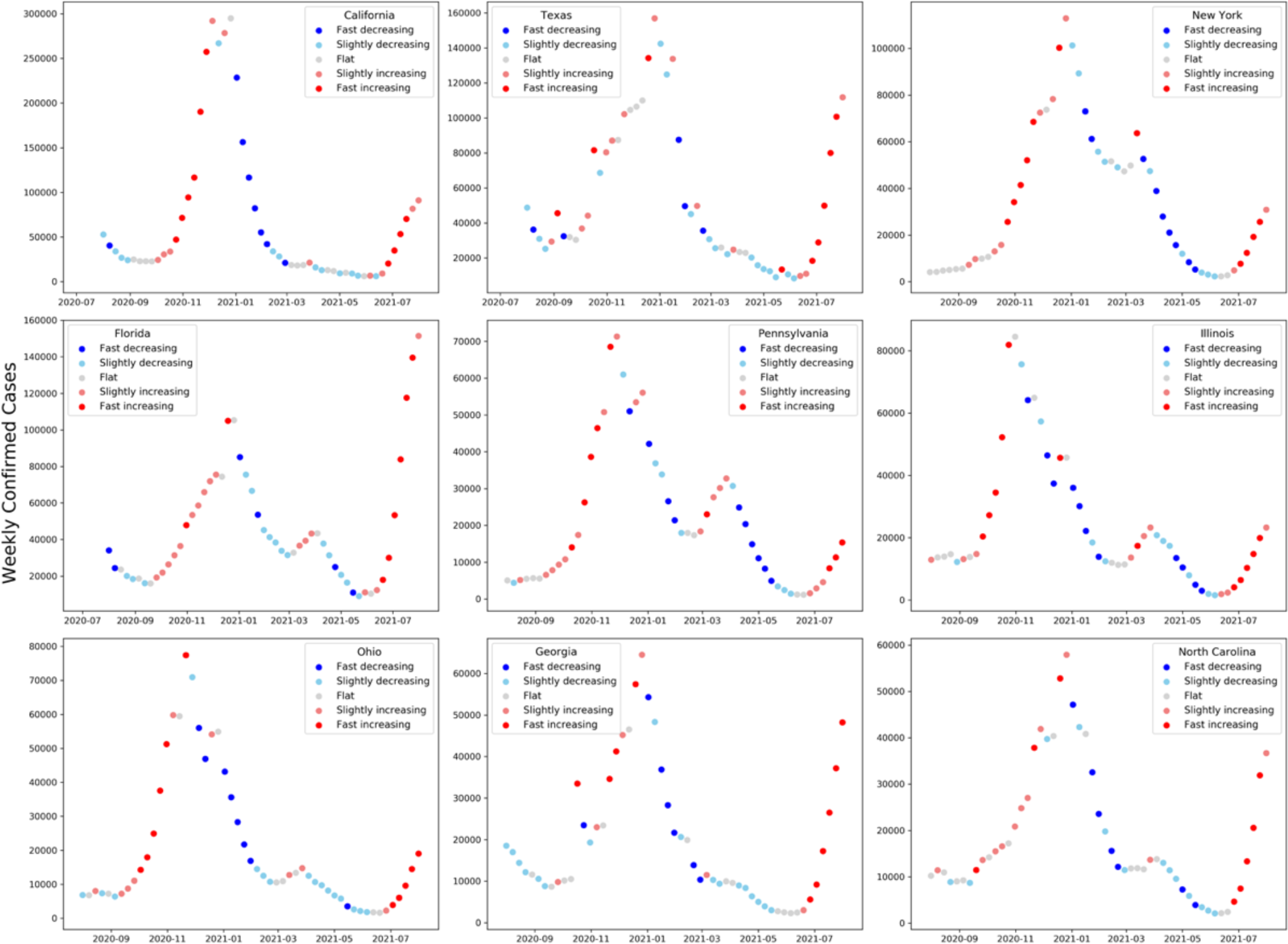
Week group based on weekly growth rate for nine selected states. The y-axis represents the weekly reported cases and the color of the dots indicates the cluster group for the given week.

#### 3.5 Model Performance by Outbreak Phase

**Supplementary Figure 16:**
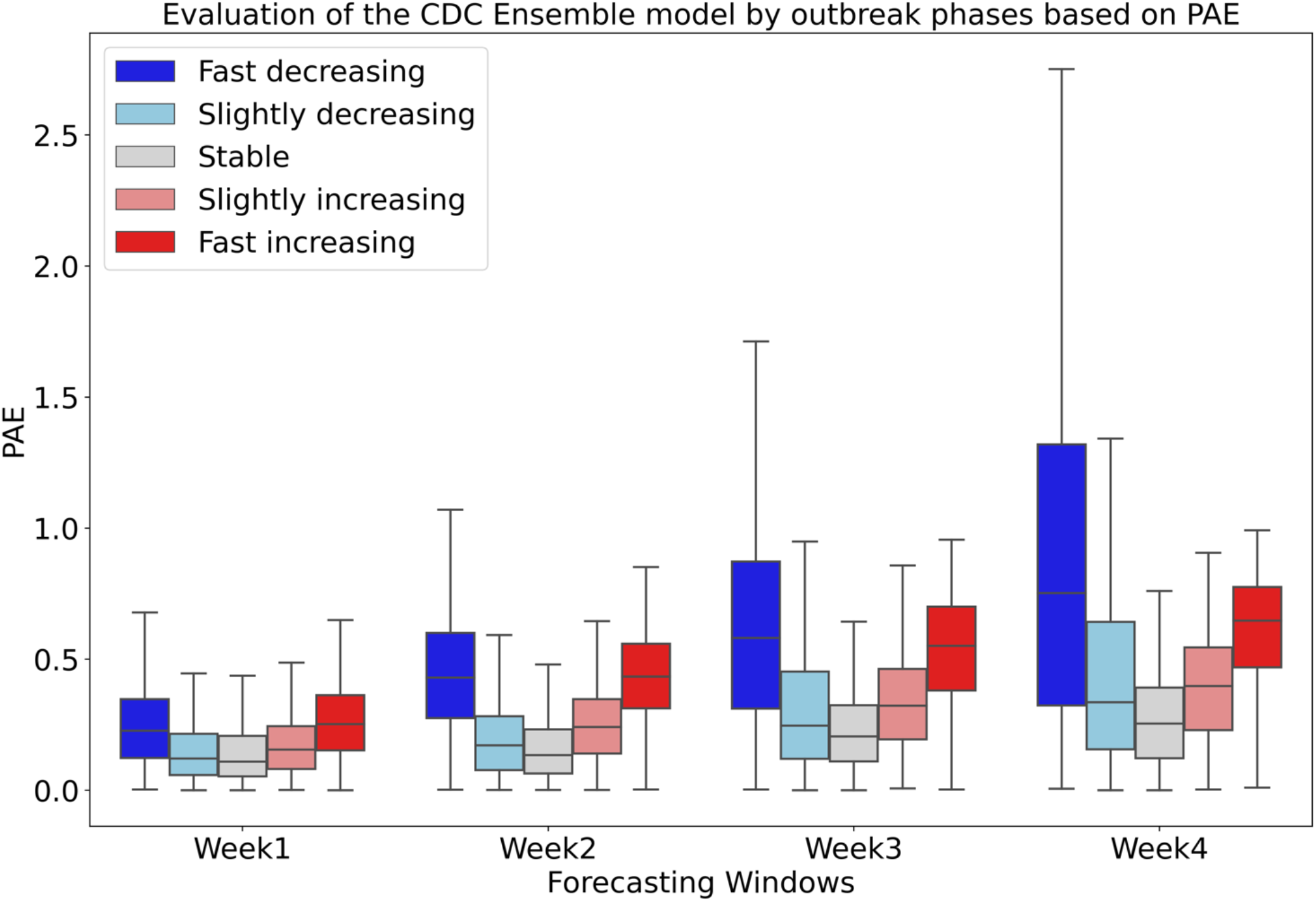
Evaluation of the CDC Ensemble model by outbreak phases based on PAE. The colors represent different outbreak phases, and each bar represents the distribution of PAE in corresponding outbreak phases.

**Supplementary Figure 17:**
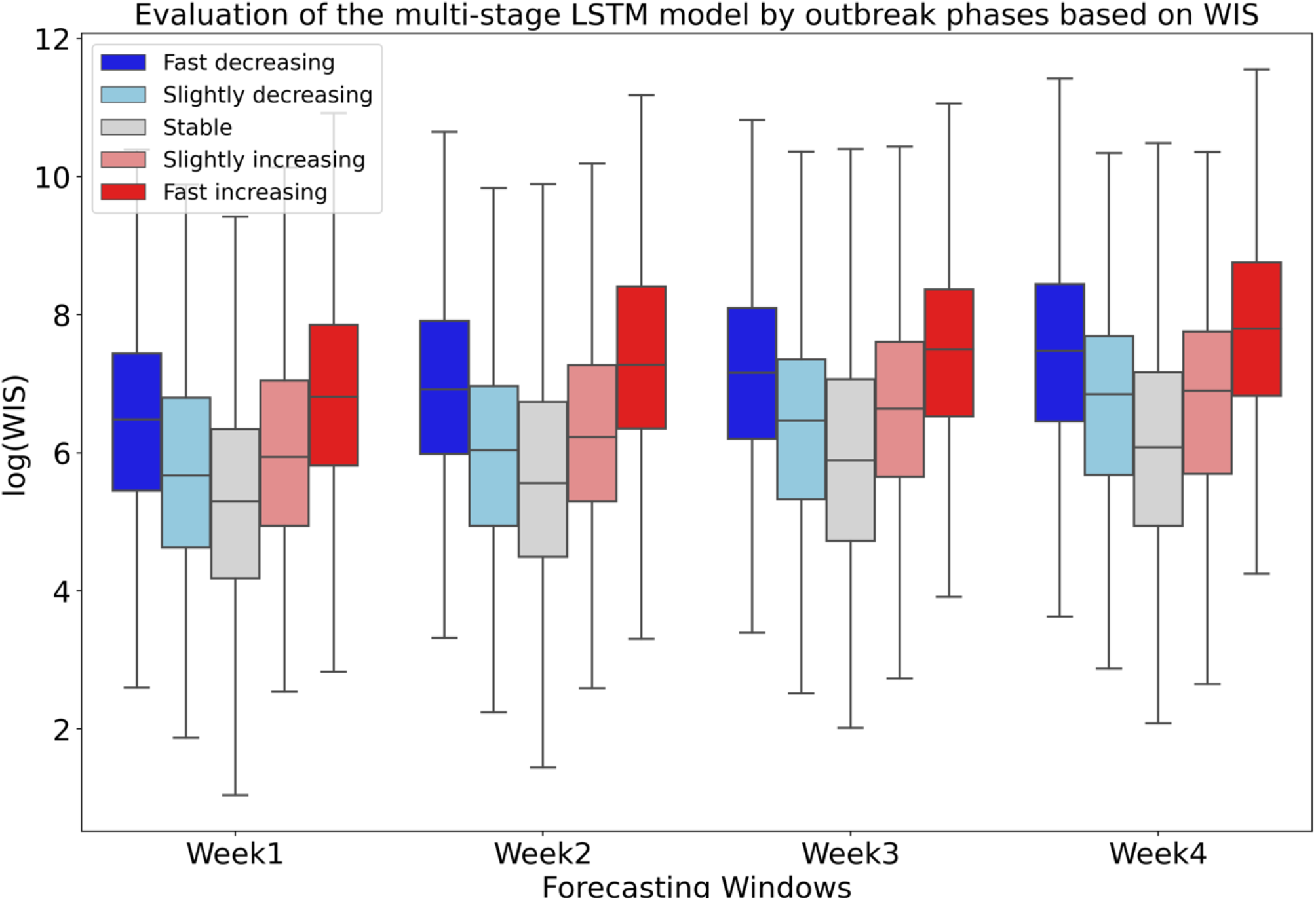
Evaluation of the multi-stage LSTM model by outbreak phases based on WIS. The colors represent different outbreak phases, and each bar represents the distribution of WIS in corresponding outbreak phases.

**Supplementary Figure 18:**
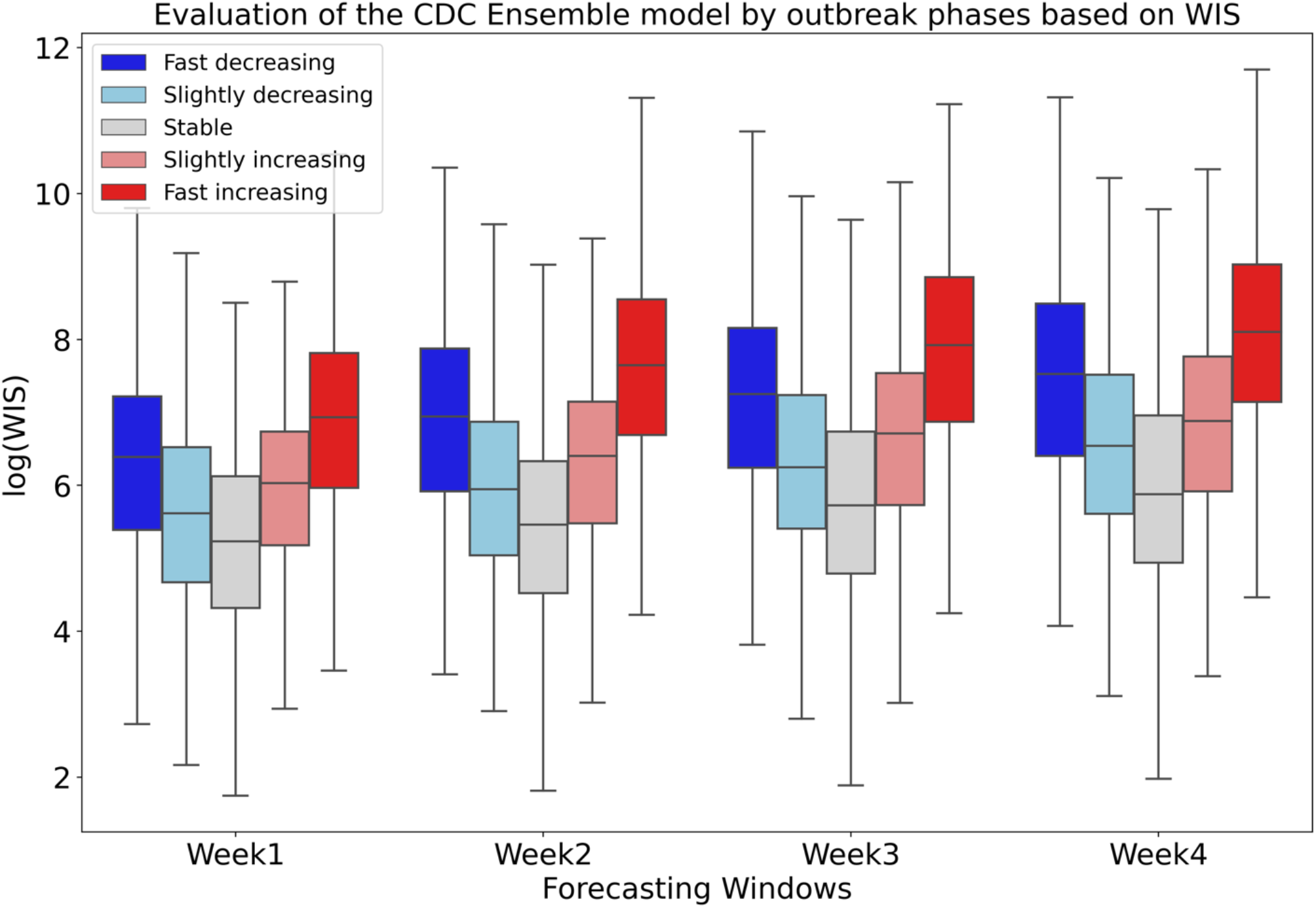
Evaluation of the CDC Ensemble model by outbreak phases based on WIS. The colors represent different outbreak phases, and each bar represents the distribution of WIS in corresponding outbreak phases.

#### 3.6 Compare model performance by outbreak with the CDC Ensemble model

In addition to phase-based evaluation of our model directly, we also quantify our model performance relative to the CDC ensemble model for each outbreak phase. For each outbreak phase group *i* and forecasting window *j*, we define a probability 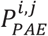 to indicate the frequency that multi-stage LSTM model outperforms the CDC ensemble model:

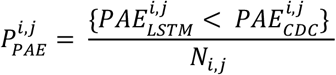

where 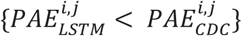 represents the number of times that the multi-stage LSTM model has smaller PAE than the CDC ensemble within phase group *i* and at forecasting window *j, N*_*i,j*_ represents the total number of predictions assigned to one of five quantile outbreak phase group *i* at forecasting window *j*. Results of 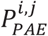 are illustrated in Appendix Figure 19, which illustrates the probability of LSTM performing better than the CDC Ensemble model based on PAE under different outbreak phases. The colors represent different outbreak phases and the y axis plots the 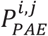. The grey dash line indicates a probability of 0·5. The results based on AE are equivalent to PAE, hence we only present the analysis based on PAE and WIS.

**Supplementary Figure 19:**
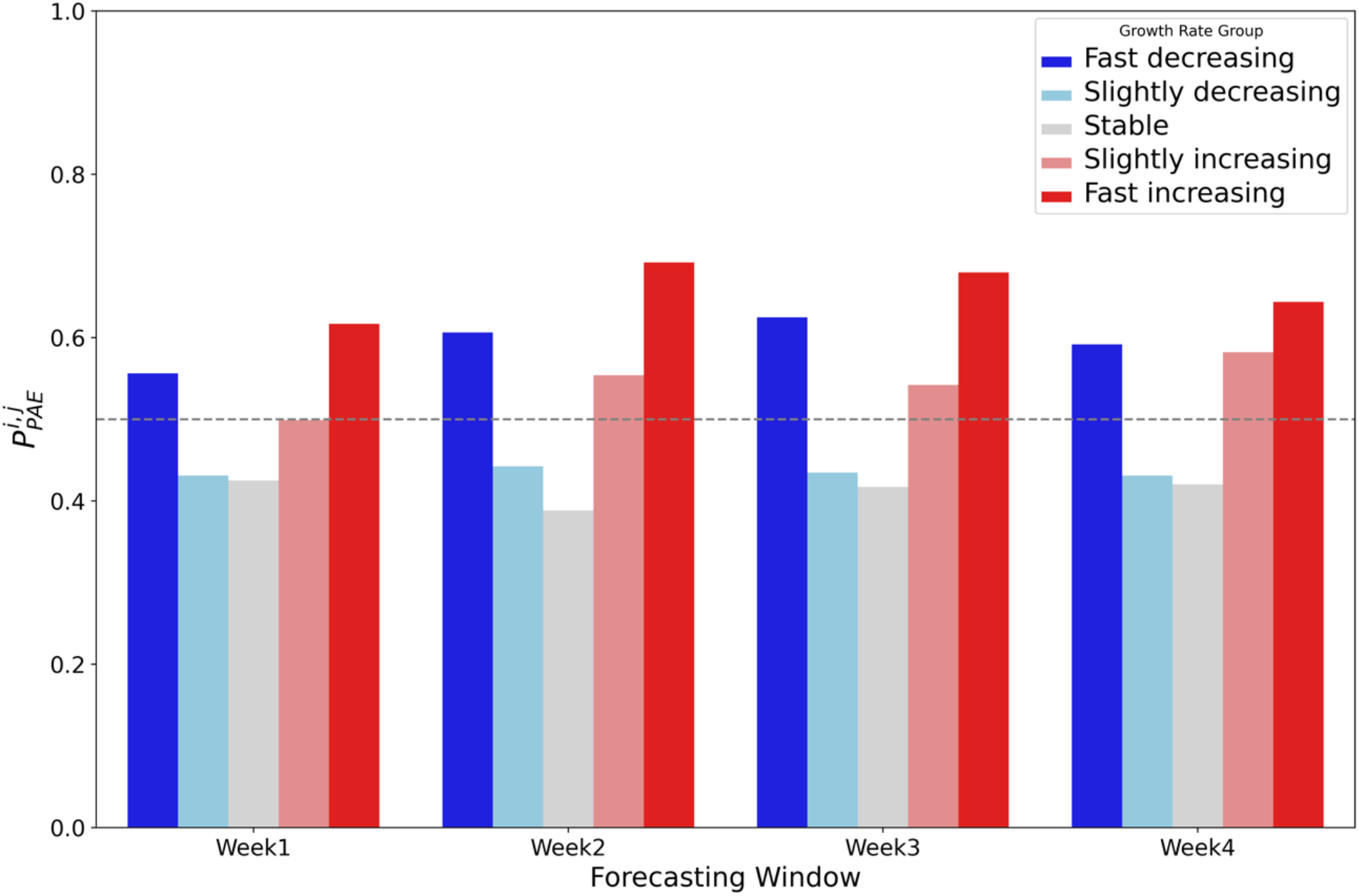
The probability of LSTM performing better than the CDC Ensemble model based on PAE under different outbreak phase. The colors represent different outbreak phase and y axis plots the probability that the multi-stage LSTM outperforms the CDC ensemble under each phase group. The grey dash line indicates a probability of 0·5.

Similar to the definition of 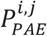, we define 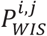 as the probability that multi-stage LSTM model outperforms the CDC ensemble model based on WIS:

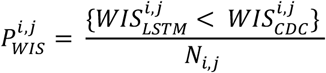

where 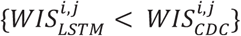 represents the number of times that the multi-stage LSTM model have smaller WIS than the CDC ensemble within phase group *i* and at forecasting window *j, N*_*i,j*_ represents the total number of predictions assigned to quantile outbreak phase group *i* at forecasting window *j*. Results of 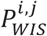 are illustrated in Appendix Figure 20, which illustrates the probability of LSTM performing better than the CDC Ensemble model based on WIS under different outbreak phases.

**Supplementary Figure 20:**
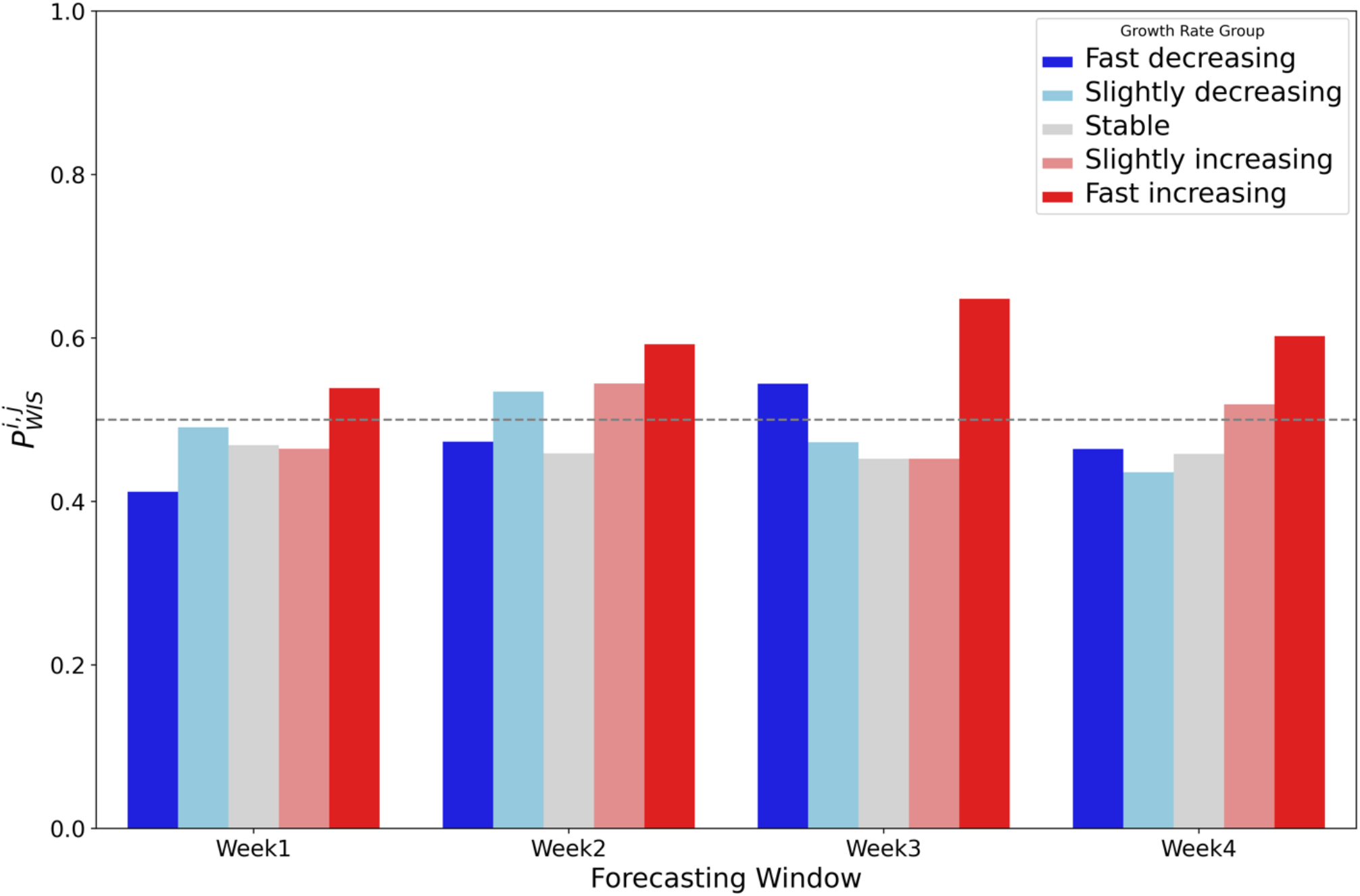
The probability of LSTM performing better than the CDC Ensemble model based on WIS under different outbreak phase. The colors represent different outbreak phase and y axis plots the probability that the multi-stage LSTM outperforms the CDC ensemble under each phase group. The grey dash line indicates a probability of 0·5.

#### 3.7 Comparing model performance after adding genomic cases data by AE and WIS

Appendix Figure 21 and 22 illustrate the results for three different models based on AE and WIS: (a) Multi-stage LSTM model without variant cases data, (b) Multi-stage LSTM model with variant cases data and (c) CDC Ensemble model. The x-axis is the week that the predictions are made on. Each pair of bar plots represents PAE distribution for the selected states at a given week, where the green bar represents the error distribution for the multi-stage LSTM model without genomic data, purple bar represents the error distribution for the multi-stage LSTM model with genomic data, and the yellow bar represents the error distribution for the CDC ensemble model.

**Supplementary Figure 21:**
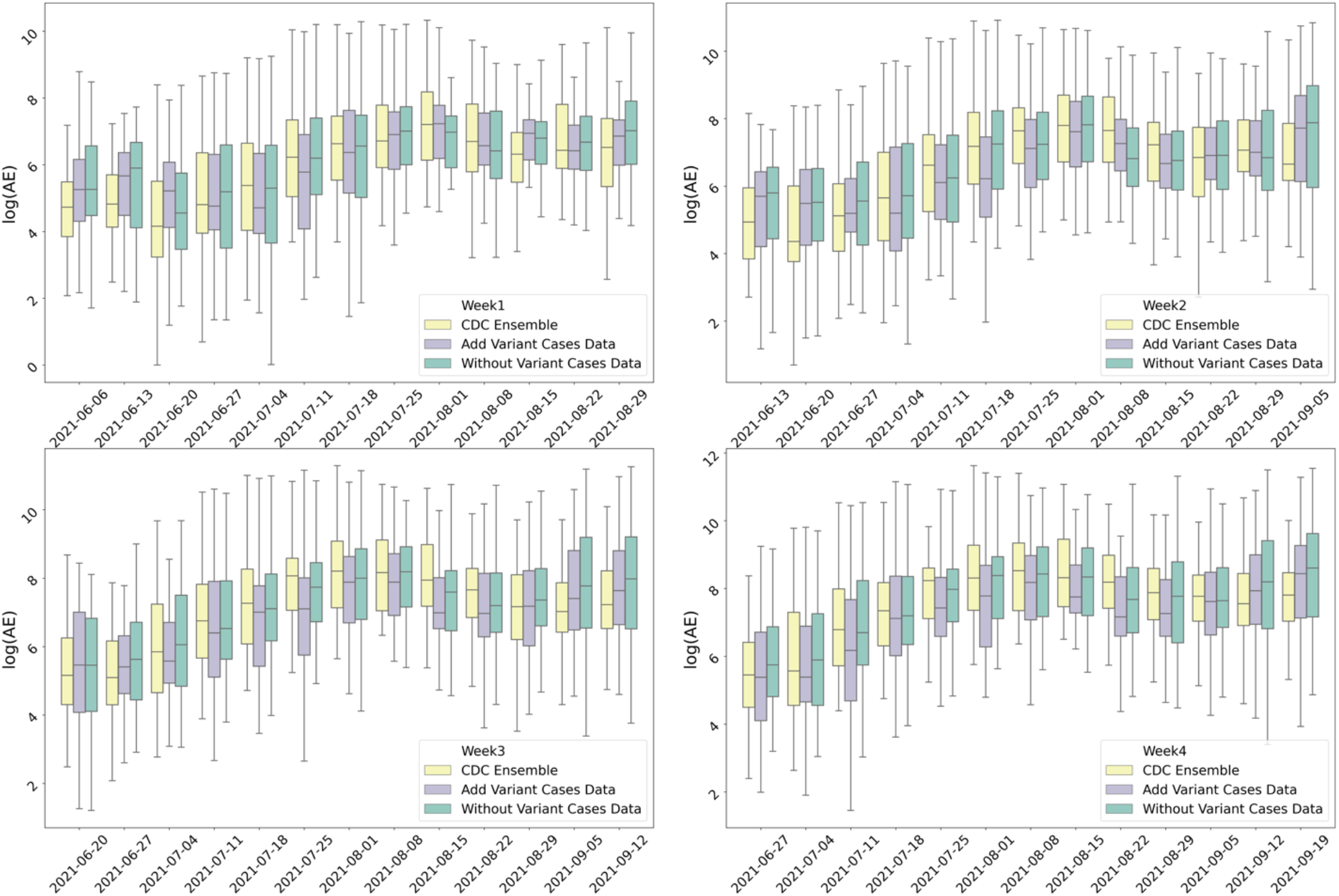
Model performance based on log(AE). for three different models: (a) Multi-stage LSTM model without variant cases data, (b) Multi-stage LSTM model with variant cases data and (c) CDC Ensemble model. The x-axis is the week that the predictions are made on. Each pair of bar plots represents PAE distribution for the selected states at a given week, where the green bar represents the error distribution for the multi-stage LSTM model without genomic data, purple bar represents the error distribution for the multi-stage LSTM model with genomic data, and the yellow bar represents the error distribution for the CDC ensemble model.

**Supplementary Figure 22:**
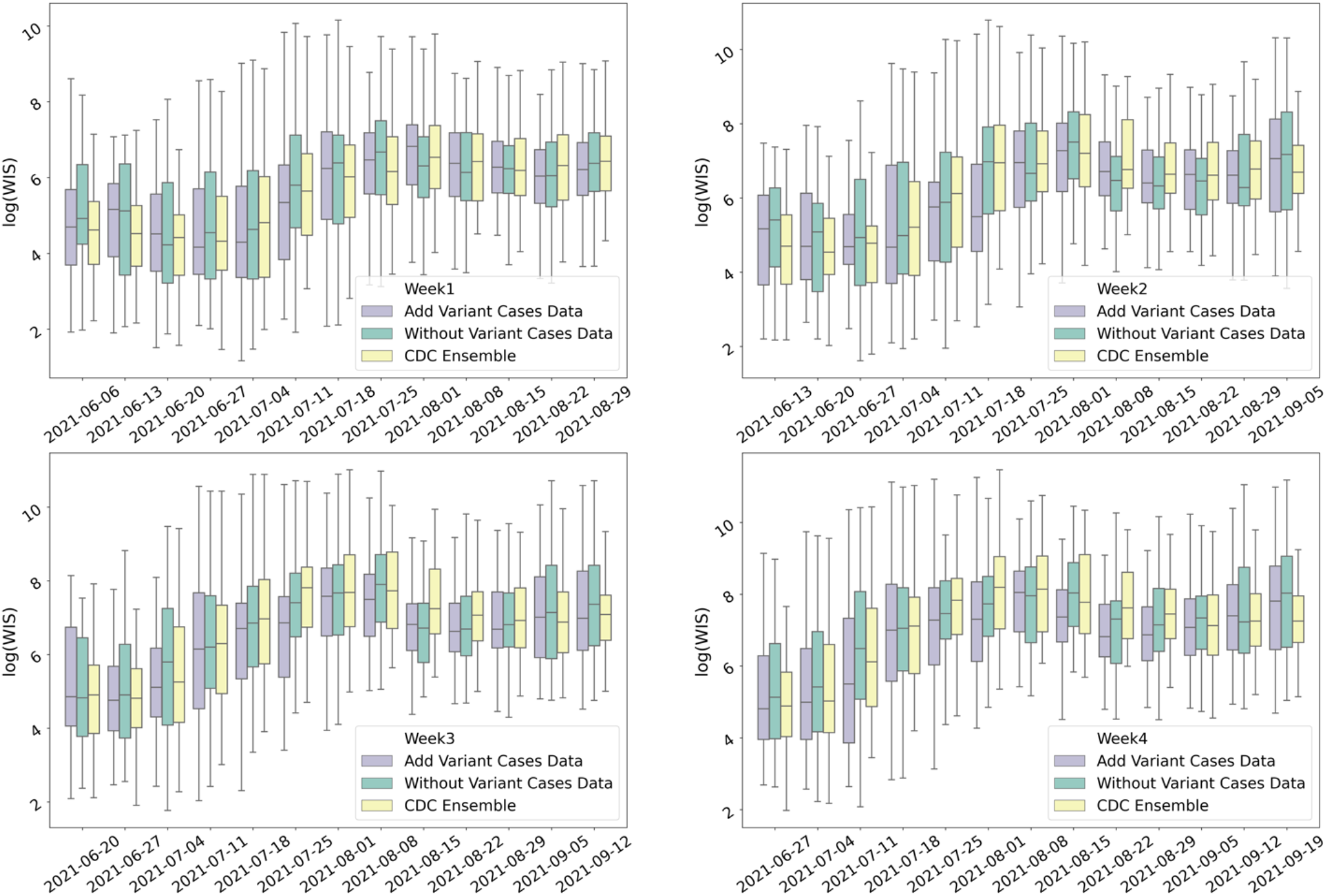
Model performance based on log(WIS). for three different models: (a) Multi-stage LSTM model without variant cases data, (b) Multi-stage LSTM model with variant cases data and (c) CDC Ensemble model. The x-axis is the week that the predictions are made on. Each pair of bar plots represents PAE distribution for the selected states at a given week, where the green bar represents the error distribution for the multi-stage LSTM model without genomic data, purple bar represents the error distribution for the multi-stage LSTM model with genomic data, and the yellow bar represents the error distribution for the CDC ensemble model.

#### 3.8 Results of weekly deaths forecasting

We also apply the models for all epidemiological weeks from September 2020 to September 2021, each week we make weekly death predictions for the next 4 weeks. Appendix Figure 23-25 compares the model performance of the Multi-stage LSTM model with the CDC ensemble model at the state-level based on different error metric. Each pair of bar plots represents error distribution for all the states at given week, where the green bar represents the error distribution for the Multi-stage LSTM model and the yellow bar represents the error distribution for the CDC ensemble model. The red curve indicates the weekly reported deaths at the national level.

**Supplementary Figure 23:**
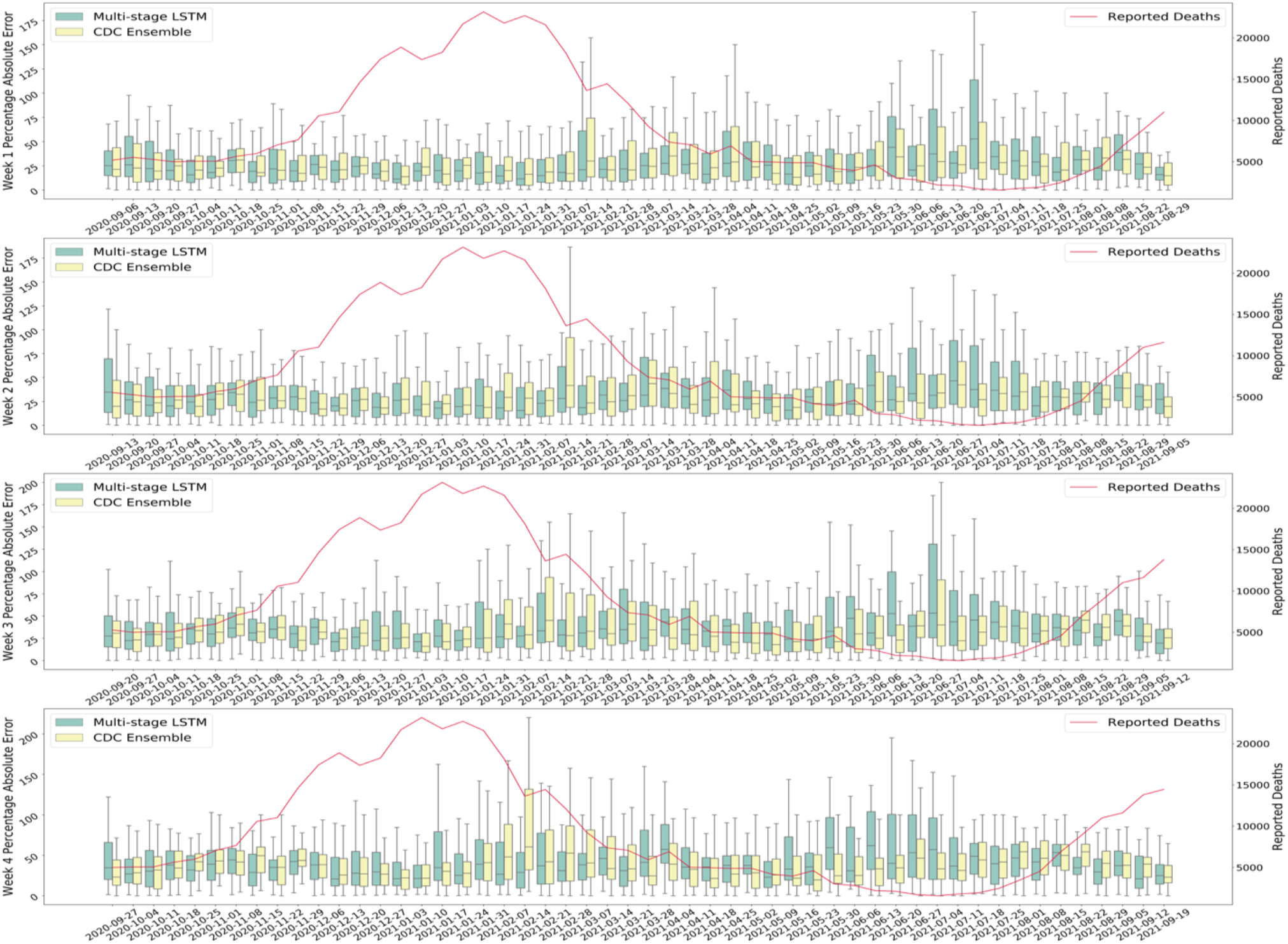
Comparison of Deaths performance between the multi-stage LSTM Model and the CDC ensemble model based on PAE. The y-axis represents the PAE for 1-4 weeks deaths’ prediction results. Each pair of bar plots represents PAE distribution for all the states at a given week, where the green bar represents the error distribution for the multi-stage LSTM model, and the yellow bar represents the error distribution for the CDC ensemble model. The red curve represents the weekly reported deaths at the national level. The left y-axis represents the PAE by different forecasting windows and right y-axis represents national level reported deaths.

**Supplementary Figure 24:**
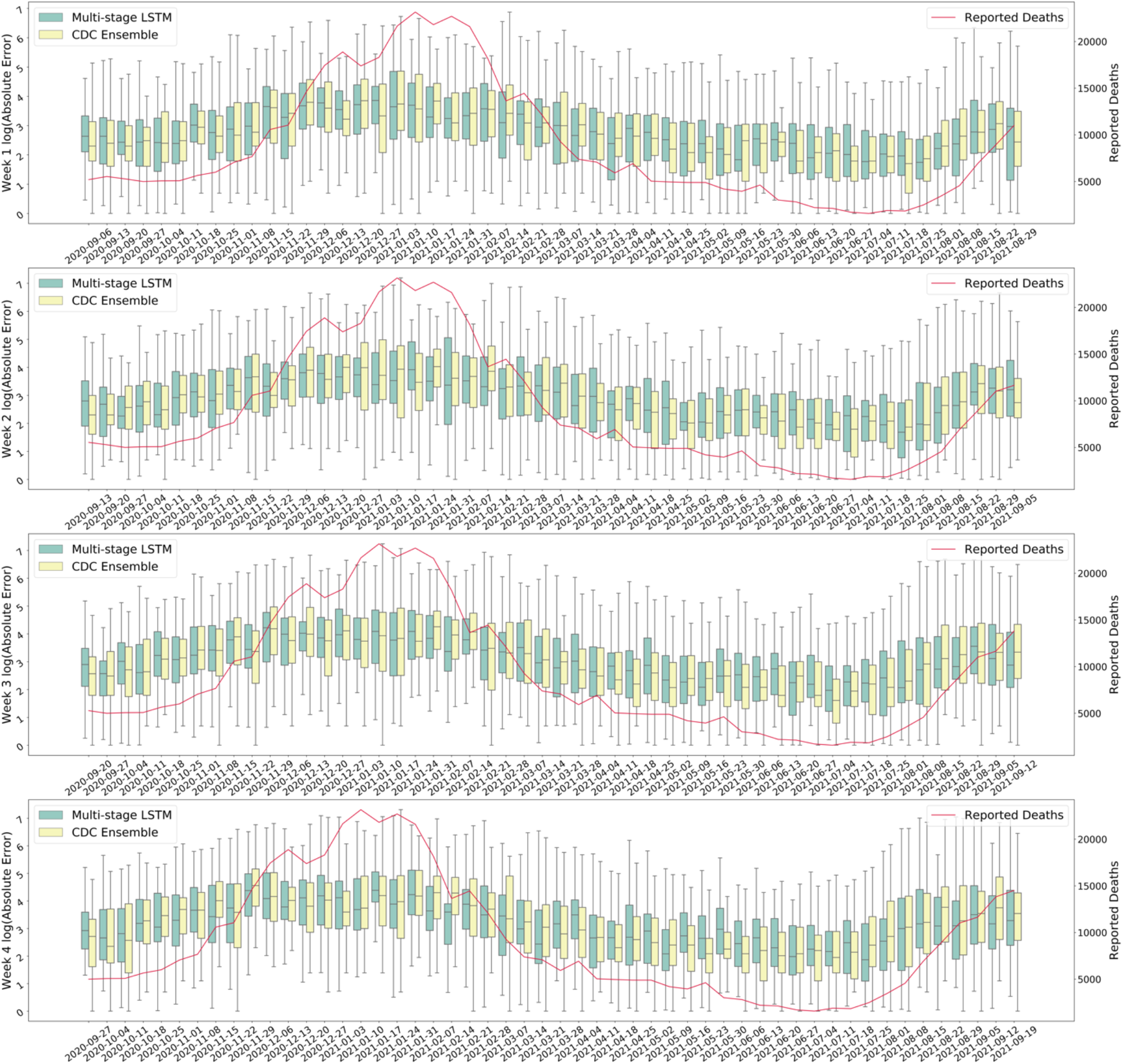
Comparison of Deaths performance between the multi-stage LSTM Model and the CDC ensemble model based on log AE. The y-axis represents the AE for 1-4 weeks deaths’ prediction results. The y-axis represents the log(AE) for 1-4 weeks deaths’ prediction results. Each pair of bar plots represents log(AE) distribution for all the states at a given week, where the green bar represents the error distribution for the multi-stage LSTM model, and the yellow bar represents the error distribution for the CDC ensemble model. The red curve represents the weekly reported deaths at the national level. The left y-axis represents the log(AE) by different forecasting windows and right y-axis represents national level reported deaths.

**Supplementary Figure 25:**
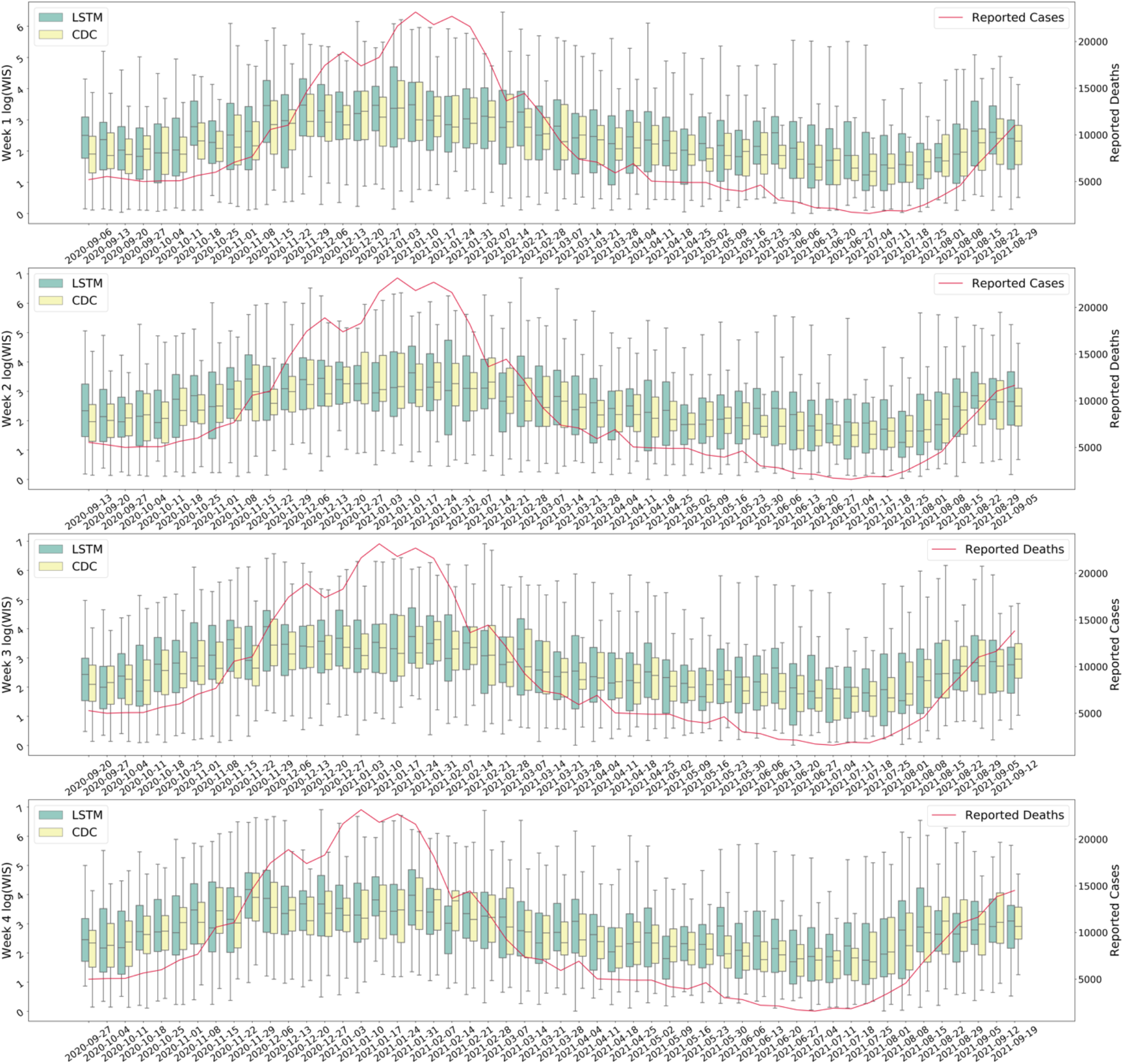
Comparison of Deaths performance between the multi-stage LSTM Model and the CDC ensemble model based on log WIS. The y-axis represents the log WIS for 1-4 weeks deaths’ prediction results. The y-axis represents the log(WIS) for 1-4 weeks deaths’ prediction results. Each pair of bar plots represents log(WIS) distribution for all the states at a given week, where the green bar represents the error distribution for the multi-stage LSTM model, and the yellow bar represents the error distribution for the CDC ensemble model. The red curve represents the weekly reported deaths at the national level. The left y-axis represents the log(WIS) by different forecasting windows and right y-axis represents national level reported deaths.

For model selection for deaths prediction model, same as cases prediction model, we assign data into four categories and add hospitalization as part of the epidemiological data. The model performance based on different input data is shown in Appendix Figure 26.

**Supplementary Figure 26:**
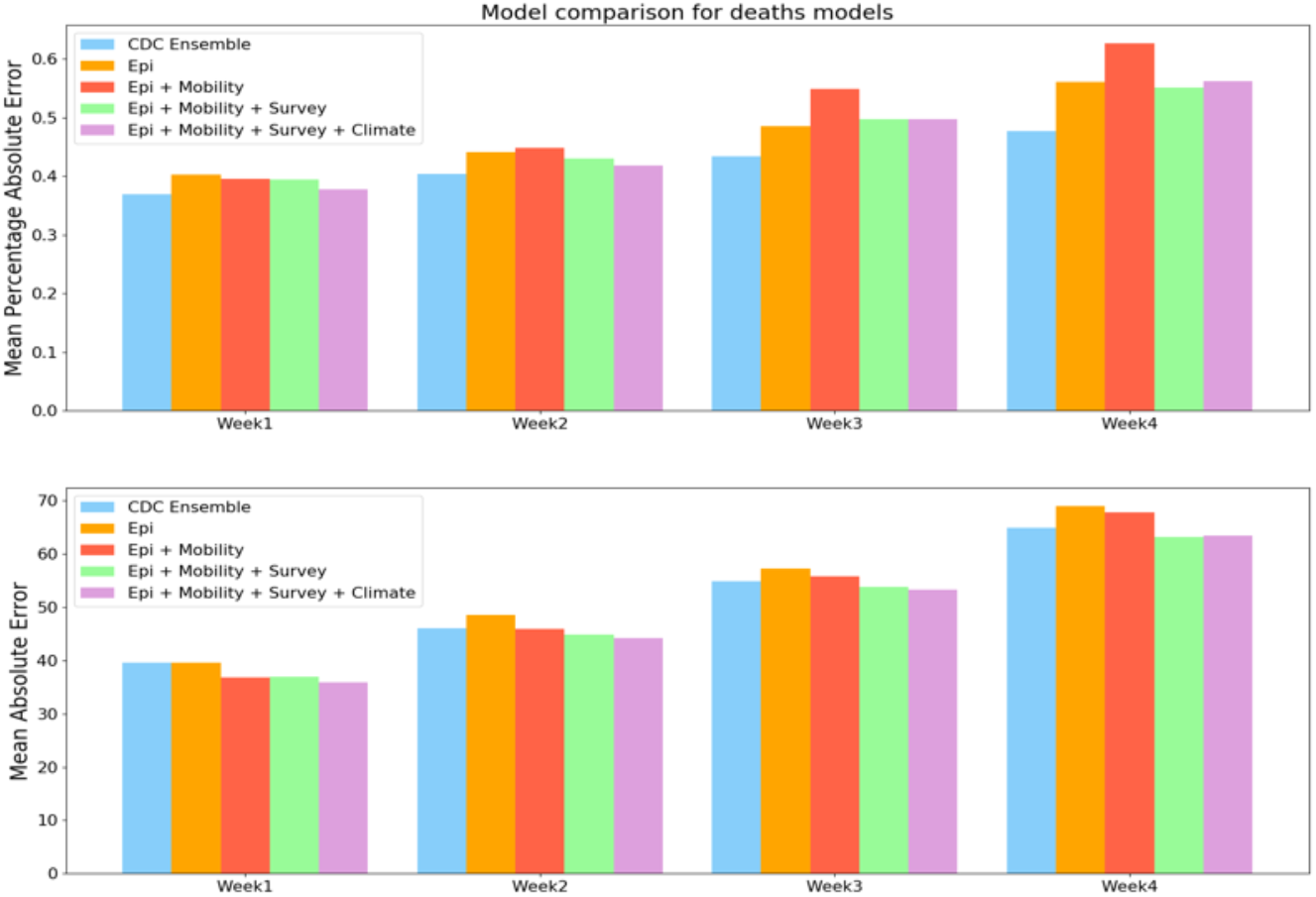
Model comparison for deaths models by mean PAE and mean AE for the period between September 2020 to September 2021. The y-axis represents the average absolute error for 1-4 weeks deaths’ prediction results across selected period.

## Notes

### Competing Interest Statement

The authors have declared no competing interest.

## References

1 Dong E, Du H, Gardner L. An interactive web-based dashboard to track COVID-19 in real time. The Lancet Infectious Diseases 2020; 20: 533–4.

2 CDC. CDC Museum COVID-19 Timeline. Centers for Disease Control and Prevention. 2021; published online Aug 4. https://www.cdc.gov/museum/timeline/covid19.html (accessed Jan 1, 2022).

3 Fanelli D, Piazza F. Analysis and forecast of COVID-19 spreading in China, Italy and France. Chaos, Solitons & Fractals 2020; 134: 109761.

4 Dansana D, Kumar R, Parida A, et al. Using Susceptible-Exposed-Infectious-Recovered Model to Forecast Coronavirus Outbreak. Cmc-Computers Materials & Continua 2021; : 1595–612.

5 Petropoulos F, Makridakis S. Forecasting the novel coronavirus COVID-19. PLOS ONE 2020; 15: e0231236.

6 Paiva HM, Afonso RJM, Oliveira IL de, Garcia GF. A data-driven model to describe and forecast the dynamics of COVID-19 transmission. PLOS ONE 2020; 15: e0236386.

7 Yang C, Wang J. Modeling the transmission of COVID-19 in the US – A case study. Infectious Disease Modelling 2021; 6: 195–211.

8 Tsay C, Lejarza F, Stadtherr MA, Baldea M. Modeling, state estimation, and optimal control for the US COVID-19 outbreak. Sci Rep 2020; 10: 10711.

9 Reiner RC, Barber RM, Collins JK, et al. Modeling COVID-19 scenarios for the United States. Nat Med 2021; 27: 94–105.

10 Zhou Y, Wang L, Zhang L, et al. A Spatiotemporal Epidemiological Prediction Model to Inform County-Level COVID-19 Risk in the United States. Harvard Data Science Review 2020; published online Aug 6. DOI:10.1162/99608f92.79e1f45e.

11 Lynch CJ, Gore R. Short-Range Forecasting of COVID-19 During Early Onset at County, Health District, and State Geographic Levels Using Seven Methods: Comparative Forecasting Study. J Med Internet Res 2021; 23: e24925.

12 Desai PS. News Sentiment Informed Time-series Analyzing AI (SITALA) to curb the spread of COVID-19 in Houston. Expert Systems with Applications 2021; 180: 115104.

13 Watson GL, Xiong D, Zhang L, et al. Pandemic velocity: Forecasting COVID-19 in the US with a machine learning & Bayesian time series compartmental model. PLOS Computational Biology 2021; 17: e1008837.

14 Zhang-James Y, Hess J, Salekin A, et al. A seq2seq model to forecast the COVID-19 cases, deaths and reproductive R numbers in US counties. 2021.

15 Ramchandani A, Fan C, Mostafavi A. DeepCOVIDNet: An Interpretable Deep Learning Model for Predictive Surveillance of COVID-19 Using Heterogeneous Features and Their Interactions. IEEE Access 2020; 8: 159915–30.

16 Nikparvar B, Rahman MM, Hatami F, Thill J-C. Spatio-temporal prediction of the COVID-19 pandemic in US counties: modeling with a deep LSTM neural network. Sci Rep 2021; 11: 21715.

17 Luo J, Zhang Z, Fu Y, Rao F. Time series prediction of COVID-19 transmission in America using LSTM and XGBoost algorithms. Results in Physics 2021; 27: 104462.

18 Shastri S, Singh K, Kumar S, Kour P, Mansotra V. Time series forecasting of Covid-19 using deep learning models: India-USA comparative case study. Chaos, Solitons & Fractals 2020; 140: 110227.

19 Fox SJ, Lachmann M, Tec M, et al. Real-time pandemic surveillance using hospital admissions and mobility data. Proceedings of the National Academy of Sciences 2022; 119: e2111870119.

20 Ray EL, Wattanachit N, Niemi J, et al. Ensemble Forecasts of Coronavirus Disease 2019 (COVID-19) in the U.S. Epidemiology, 2020 DOI:10.1101/2020.08.19.20177493.

21 Wu D, Gao L, Xiong X, et al. DeepGLEAM: A hybrid mechanistic and deep learning model for COVID-19 forecasting. arXiv:210206684 [cs] 2021; published online March 23. http://arxiv.org/abs/2102.06684 (accessed Dec 6, 2021).

22 Cramer EY, Ray EL, Lopez VK, et al. Evaluation of individual and ensemble probabilistic forecasts of COVID-19 mortality in the US. 2021; : 2021.02.03.21250974.

23 Nixon K, Jindal S, Parker F, et al. An Evaluation of Prospective COVID-19 Modeling: From Data to Science Translation. 2022; : 2022.04.18.22273992.

24 Pollett S, Johansson MA, Reich NG, et al. Recommended reporting items for epidemic forecasting and prediction research: The EPIFORGE 2020 guidelines. PLOS Medicine 2021; 18: e1003793.

25 Cramer EY, Huang Y, Wang Y, et al. The United States COVID-19 Forecast Hub dataset. 2021.

26 Home - COVID 19 forecast hub. https://covid19forecasthub.org/ (accessed March 21, 2022).

27 Earnest R, Uddin R, Matluk N, et al. Comparative transmissibility of SARS-CoV-2 variants Delta and Alpha in New England, USA. Cell Reports Medicine 2022; 3: 100583.

28 Kalia K, Saberwal G, Sharma G. The lag in SARS-CoV-2 genome submissions to GISAID. Nat Biotechnol 2021; 39: 1058–60.

29 Sera F, Armstrong B, Abbott S, et al. A cross-sectional analysis of meteorological factors and SARS-CoV-2 transmission in 409 cities across 26 countries. Nat Commun 2021; 12: 5968.

30 U.S. Department of Health & Human Services (HHS). HHS.gov. https://www.hhs.gov/index.html (accessed March 21, 2022).

31 David C. Farrow, Logan C. Brooks, Ryan J. Tibshirani, Roni Rosenfeld. Delphi Epidata API. GitHub. https://github.com/cmu-delphi/delphi-epidata (accessed March 21, 2022).

32 Places Data & Foot Traffic Insights | SafeGraph. https://www.safegraph.com/ (accessed Oct 22, 2021).

33 Badr HS, Zaitchik BF, Kerr GH, et al. Unified real-time environmental-epidemiological data for multiscale modeling of the COVID-19 pandemic. Epidemiology, 2021 DOI:10.1101/2021.05.05.21256712.

34 Bureau UC. State Population by Characteristics: 2010-2019. Census.gov. https://www.census.gov/data/tables/time-series/demo/popest/2010s-state-detail.html (accessed March 21, 2022).

35 Elbe S, Buckland-Merrett G. Data, disease and diplomacy: GISAID’s innovative contribution to global health. Global Challenges 2017; 1: 33–46.

## Reference

1 Friedman J, Liu P, Troeger CE, et al. Predictive performance of international COVID-19 mortality forecasting models. Nature communications 2021; 12: 1–13.

2 Vaid S, Cakan C, Bhandari M. Using machine learning to estimate unobserved COVID-19 infections in North America. The Journal of bone and joint surgery American volume 2020.

3 Anastassopoulou C, Russo L, Tsakris A, Siettos C. Data-based analysis, modelling and forecasting of the COVID-19 outbreak. PloS one 2020; 15: e0230405.

4 Dong E, Du H, Gardner L. An interactive web-based dashboard to track COVID-19 in real time. The Lancet Infectious Diseases 2020; 20: 533–4.

5 COVID-19 data. Centers for Civic Impact, 2022 https://github.com/govex/COVID-19 (accessed June 14, 2022).

6 Dialysis COVID-19 Vaccination Data Dashboard | NHSN | CDC. 2022; published online Jan 31. https://www.cdc.gov/nhsn/covid19/dial-vaccination-dashboard.html (accessed June 14, 2022).

7 U.S. Department of Health & Human Services (HHS). HHS.gov. https://www.hhs.gov/index.html (accessed March 21, 2022).

8 David C. Farrow, Logan C. Brooks, Ryan J. Tibshirani, Roni Rosenfeld. Delphi Epidata API. GitHub. 2015. https://github.com/cmu-delphi/delphi-epidata (accessed March 21, 2022).

9 Badr HS, D. H, Marshall M, Dong E, Squire MM, Gardner LM. Association between mobility patterns and COVID-19 transmission in the USA: a mathematical modelling study. The Lancet Infectious Diseases 2020; 20: 1247–54.

10 Ilin C, Annan-Phan S, Tai XH, Mehra S, Hsiang S, Blumenstock JE. Public mobility data enables COVID-19 forecasting and management at local and global scales. Sci Rep 2021; 11: 13531.

11 Chang S, Pierson E, Koh PW, et al. Mobility network models of COVID-19 explain inequities and inform reopening. Nature 2021; 589: 82–7.

12 Guan G, Dery Y, Yechezkel M, Ben-Gal I, Yamin D, Brandeau ML. Early detection of COVID-19 outbreaks using human mobility data. PLOS ONE 2021; 16: e0253865.

13 Limitations of using mobile phone data to model COVID-19 transmission in the USA - The Lancet Infectious Diseases. https://www.thelancet.com/journals/laninf/article/PIIS1473-3099(20)30861-6/fulltext (accessed March 22, 2022).

14 Gatalo O, Tseng K, Hamilton A, Lin G, Klein E. Associations between phone mobility data and COVID-19 cases. The Lancet Infectious Diseases 2021; 21: e111.

15 Places Data Curated for Accurate Geospatial Analytics | SafeGraph. https://www.safegraph.com (accessed Oct 14, 2021).

16 Weekly Patterns | SafeGraph Docs. SafeGraph. https://docs.safegraph.com/docs/weekly-patterns (accessed March 22, 2022).

17 Core Places | SafeGraph Docs. https://docs.safegraph.com/docs/core-places (accessed March 22, 2022).

18 Pearson K. LIII. On lines and planes of closest fit to systems of points in space. The London, Edinburgh, and Dublin philosophical magazine and journal of science 1901; 2: 559–72.

19 McDonald D, Bien J, Green A, Hu AJ, Tibshirani R. Replication Data for: Can Auxiliary Indicators Improve COVID-19 Forecasting and Hotspot Prediction? 2021; published online Nov 8. DOI:10.5683/SP3/UW4VTC.

20 Reinhart A, Brooks L, Jahja M, et al. An open repository of real-time COVID-19 indicators. PNAS 2021; 118. DOI:10.1073/pnas.2111452118.

21 Kerr GH, Badr HS, Gardner LM, Perez-Saez J, Zaitchik BF. Associations between meteorology and COVID-19 in early studies: Inconsistencies, uncertainties, and recommendations. One Health 2021; 12: 100225.

22 Malki Z, Atlam E-S, Hassanien AE, Dagnew G, Elhosseini MA, Gad I. Association between weather data and COVID-19 pandemic predicting mortality rate: Machine learning approaches. Chaos, Solitons & Fractals 2020; 138: 110137.

23 Sera F, Armstrong B, Abbott S, et al. A cross-sectional analysis of meteorological factors and SARS-CoV-2 transmission in 409 cities across 26 countries. Nat Commun 2021; 12: 5968.

24 Badr HS, Zaitchik BF, Kerr GH, et al. Unified real-time environmental-epidemiological data for multiscale modeling of the COVID-19 pandemic. medRxiv 2021.

25 Dowd JB, Andriano L, Brazel DM, et al. Demographic science aids in understanding the spread and fatality rates of COVID-19. Proc Natl Acad Sci U S A 2020; 117: 9696–8.

26 Bureau UC. State Population by Characteristics: 2010-2019. Census.gov. https://www.census.gov/data/tables/time-series/demo/popest/2010s-state-detail.html (accessed March 21, 2022).

27 Elbe S, Buckland-Merrett G. Data, disease and diplomacy: GISAID’s innovative contribution to global health. Global challenges 2017; 1: 33–46.

28 Hochreiter S, Schmidhuber J. Long short-term memory. Neural computation 1997; 9: 1735–80.

29 Bracher J, Ray EL, Gneiting T, Reich NG. Evaluating epidemic forecasts in an interval format. PLoS computational biology 2021; 17: e1008618.

